# Increased MRI-based brain age in chronic migraine patients

**DOI:** 10.1101/2022.11.21.22282575

**Authors:** Rafael Navarro-González, David García-Azorín, Ángel L. Guerrero-Peral, Álvaro Planchuelo-Gómez, Santiago Aja-Fernández, Rodrigo de Luis-García

## Abstract

Neuroimaging has revealed that migraine is linked to alterations in both the structure and function of the brain. However, the relationship of these changes with aging has not been studied in detail. Here we employ the Brain Age framework to analyze migraine, by building a machine learning model that predicts age from neuroimaging data. We hypothesize that migraine patients will exhibit an increased Brain Age Gap (the difference between the predicted age and the chronological age) compared to healthy participants. We trained a machine learning model to predict Brain Age from 2,771 T1-weighted magnetic resonance imaging scans of healthy subjects. The processing pipeline included the automatic segmentation of the images, the extraction of 1,479 imaging features (both morphological and intensity-based), harmonization, feature selection and training inside a 10-fold cross-validation scheme. Separate models based only on morphological and intensity features were also trained, and all the Brain Age models were later applied to a discovery cohort composed of 247 subjects, divided into healthy controls (HC, n=82), episodic migraine (EM, n=91), and chronic migraine patients (CM, n=74). CM patients showed an increased Brain Age Gap compared to HC (4.16 vs -0.56 years, P=0.01). A smaller Brain Age Gap was found for EM patients, not reaching statistical significance (1.21 vs -0.56 years, P=0.19). No associations were found between the Brain Age Gap and headache or migraine frequency, or duration of the disease. Brain imaging features that have previously been associated with migraine were among the main drivers of the differences in the predicted age. Also, the separate analysis using only morphological or intensity-based features revealed different patterns in the Brain Age biomarker in patients with migraine. The brain-predicted age has shown to be a sensitive biomarker of CM patients and can help reveal distinct aging patterns in migraine.

## Introduction

Migraine is a prevalent and chronic condition known for its recurrent and debilitating headache episodes. Migraine can be classified into two categories based on the frequency of headache days per month, namely episodic migraine (EM) and chronic migraine (CM) (1). Due to the inherent characteristics of migraine and its widespread occurrence, it imposes a substantial burden on both individuals and society as a whole (2).

Migraine is associated with changes in the brain. Beyond the direct effects (i.e., the experience of pain during the ictal phase), neuroimaging studies have discovered alterations in the migrainous brain during the interictal phase encompassing both the structural and functional levels (3–5). Differences between EM and CM have also been reported (6–8). Even though the structure and function of the brain are also impacted by changes due to brain development and aging, the interplay between those and changes related to migraine has not been explored in depth. Bell et al. (9), for instance, focused on the pediatric age range, finding age-and puberty-dependent alterations in the functional connectivity of multiple networks in children with migraine using resting-state functional Magnetic Resonance Imaging (fMRI) and showing that brain changes associated with migraine begin in infancy and are modulated by development. Chong et al. (10) studied morphological changes of EM patients along age and found that patients with migraine have age-related thinning of regions compared to the control group. Using fluorodeoxyglucose positron emission tomography (FGD-PET), M. Lisicki et al. (11) showed that episodic migraine patients exhibit specific metabolic brain modifications while aging.

Recently, the so-called Brain Age paradigm has been proposed to explore the relationship between aging and disease (12). Using machine learning techniques from neuroimaging data, chronological age can be accurately predicted in healthy individuals. After training a Brain Age model, the difference between an individual’s chronological age and the age predicted by the Brain Age model is usually referred to as “Brain Age Gap”, “Brain Age Gap Estimate” or “brain-predicted age difference” (brain-PAD), and has been proposed as an age-adjusted index of structural brain health. Research has shown the Brain Age paradigm to be sensitive to many neurological, psychiatric, and metabolic disorders, showing a positive Brain Age Gap, higher age compared-to the healthy brain, in disorders such as Alzheimer’s, schizophrenia, and type II diabetes, among others (13–15). Conversely, protective sociological and lifestyle factors including years of education, physical exercise, playing music, or meditation have been associated with a negative Brain Age Gap (16–18). An increased predicted Brain Age has even been associated with higher allostatic load and elevated overall mortality risk (19). Even though other pain-related conditions have been studied using the Brain Age paradigm (20–23), to the best of our knowledge, migraine has not been explored from this perspective.

In this work, the Brain Age framework was employed to investigate migraine on a dataset composed of structural T1-weighted (T1w) MRI from EM and CM patients, together with normal controls. We hypothesized that migraine patients will exhibit an increased Brain Age Gap compared to healthy participants. Furthermore, we aimed to detect possible associations between the Brain Age Gap and clinical characteristics in the patient groups exploring the role of different imaging features.

## Materials and methods

Developing a robust brain age model involves several crucial steps. Firstly, it is imperative to assemble a diverse, broad, and representative dataset that encompasses neuroimaging data alongside corresponding chronological ages. The size of the dataset plays a significant role, as a larger dataset enables greater precision and generalizability in the final model. Subsequently, feature extraction is performed to capture pertinent information from the neuroimaging data. This process ensures that only informative and discriminative features are included in the model.

Once the features have been extracted, an appropriate machine learning algorithm is selected for age prediction based on the neuroimaging data. Common choices include support vector machines or neural networks. The chosen algorithm is then trained using the dataset, and techniques such as regularization, cross-validation, and hyperparameter tuning are employed to optimize performance and prevent overfitting. The trained model is next evaluated using a separate dataset, employing metrics such as mean absolute error (MAE) or correlation coefficients to assess accuracy and generalization capabilities. This evaluation step provides valuable insights into the model’s performance and its ability to accurately estimate brain age.

Finally, the trained brain age model can be applied to new and unseen neuroimaging data. In our case, we apply it to a dataset composed of healthy controls, patients with episodic migraine, and patients with chronic migraine.

### Brain age model

To create and evaluate our age prediction models, we compiled a dataset (hereinafter referred to as *Model Creation Dataset*) consisting of 2,771 structural T1w MRI scans of healthy adults aged 18 to 60 from different studies and databases that were either publicly available. These include: the Dallas Lifespan Brain Study (DLBS) (24); the Consortium for Reliability and Reproducibility dataset (CoRR) (25); the Neurocognitive aging data release (NeuroCog) (26); The OASIS-1 dataset (27); the Southwest University Adult Lifespan Dataset (SALD) (28); the Information eXtraction from Images dataset (IXI) (29); and the Cam-CAN repository (available at http://www.mrc-cbu. cam.ac.uk/datasets/camcan/) (30, 31). In addition to these, we included a set of healthy adults from the *Laboratorio de Procesado de Imagen* (LPI), our own institution. Individuals who presented neurological or psychological diagnoses or cognitive impairments were eliminated from the OASIS-1 or CoRR databases. Table 1 depicts the basic features of the *Model Creation Dataset*.

**Table 1:**
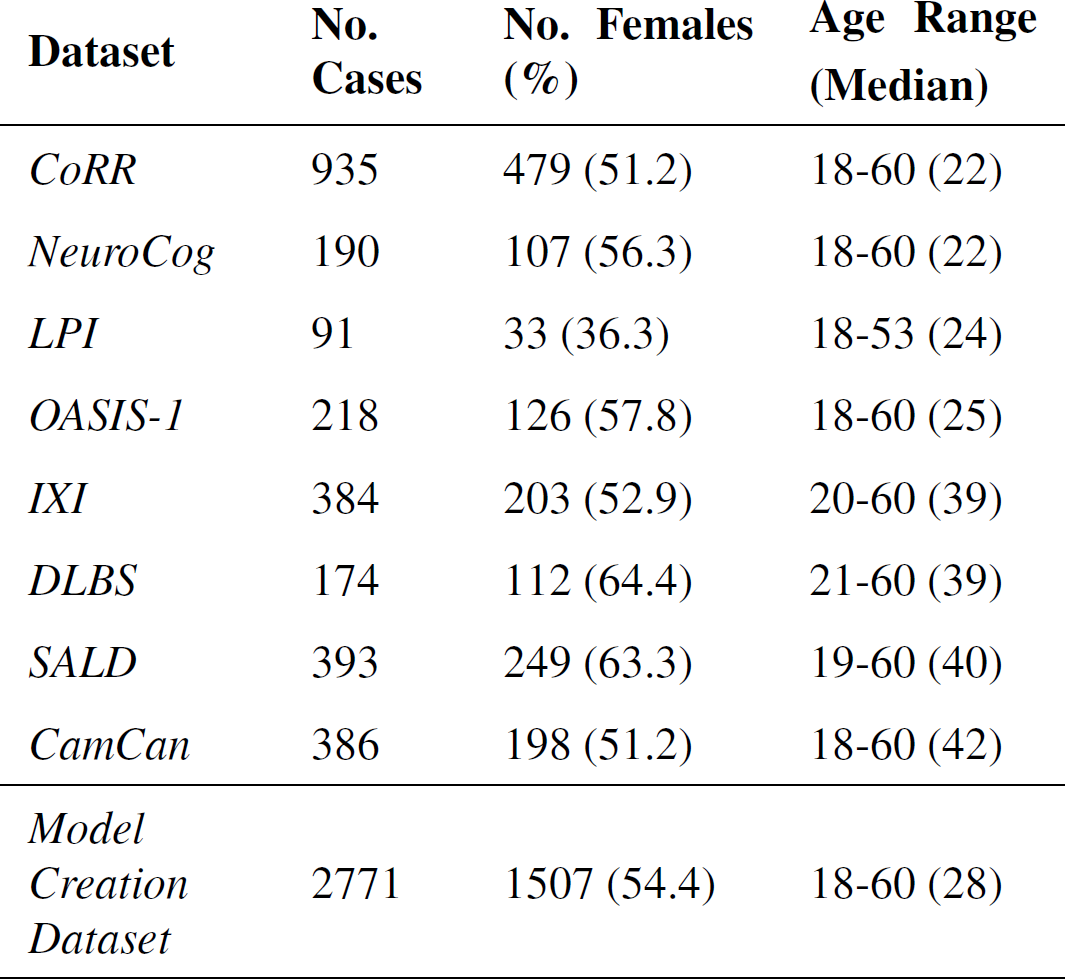
Summary characteristics of the datasets used in the *Model Creation Dataset*, sorted by median age.

From the T1w images, FastSurfer (32) was employed to extract a total of 1,479 features. Fastsurfer uses Deep Learning to perform brain segmentation based on the Desikan-Killiany atlas (33, 34). Two types of features were extracted:

- 624 morphological features, including whole brain features, the volume of cortical and subcortical gray matter regions and white matter regions from the atlas, as well as the surface, thickness and curvature of the cortical regions. This feature set will be referred to as *Morphological Feature Set*.
- 855 intensity-based features extracted from the same regions. This feature set will be referred to as *Intensity Feature Set*.

Together, all 1,479 features make up the *Combined Feature Set*. The three feature sets obtained using this procedure were the basis for further analysis.

To assure their quality, segmentations were manually inspected. In Supplementary Table 1, Supplementary Figure 1 and Supplementary Table 2, features and regions of interest are covered in greater detail.

MRI acquisitions obtained at different sites and/or using different protocols can differ in their intensity levels, which can introduce a bias in the Brain Age-predicting models. In order to cope with this problem, we used ComBatGAM (35) to harmonize the features from the *Intensity Feature Set* and the *Combined Feature Set*, using age, sex, and estimated total intracranial volume (eTIV) as covariates.

Afterwards, the cases were randomly divided into an 8:1:1 ratio for training, validation, and testing. We conducted a 10-fold cross-validation training procedure over the harmonized features to predict age. We flattened outliers of each feature, defined as values on the 97.5th or 2.5th percentile. In addition, each characteristic was adjusted to the range (-1, 1) using min-max normalization. Each fold underwent feature selection, defining three sets of 20, 30, and 40 characteristics. The selection of features was accomplished in two steps. Initially, a filter was used to choose the first decile features based on the mutual information between features and age in the training set. Next, the final feature were chosen by employing a forward feature selection approach with gaussian mixture models to optimize the mutual information between a subset of features and age (36). As regressors, support vector regressor (SVR), random forest (RF), and a multilayer perceptron (MLP) were evaluated. Figure 1 depicts the process followed. By combining these three regressors with distinct feature sets of 20, 30, and 40 characteristics for each fold, a total of 90 models were trained. Predictions were obtained for the validation and test set for each fold. Validation results were used to select the Brain Age model to be selected as the best-performing, while test results were exclusively employed to report the accuracy of the Brain Age model on the *Model Creation Dataset*. We are aware that Brain Age models suffer from regression dilution, which causes bias in Brain Age predictions. Therefore, in order to avoid possible spurious associations, a correction for this effect was applied (37, 38). A linear regression was fitted between the real age and validation results of each of the regressors of the ensemble. The intercept (*α*) and slope (*β*) of each fit were then used to correct the predictions obtained for the studied groups following the equation:

**Figure 1:**
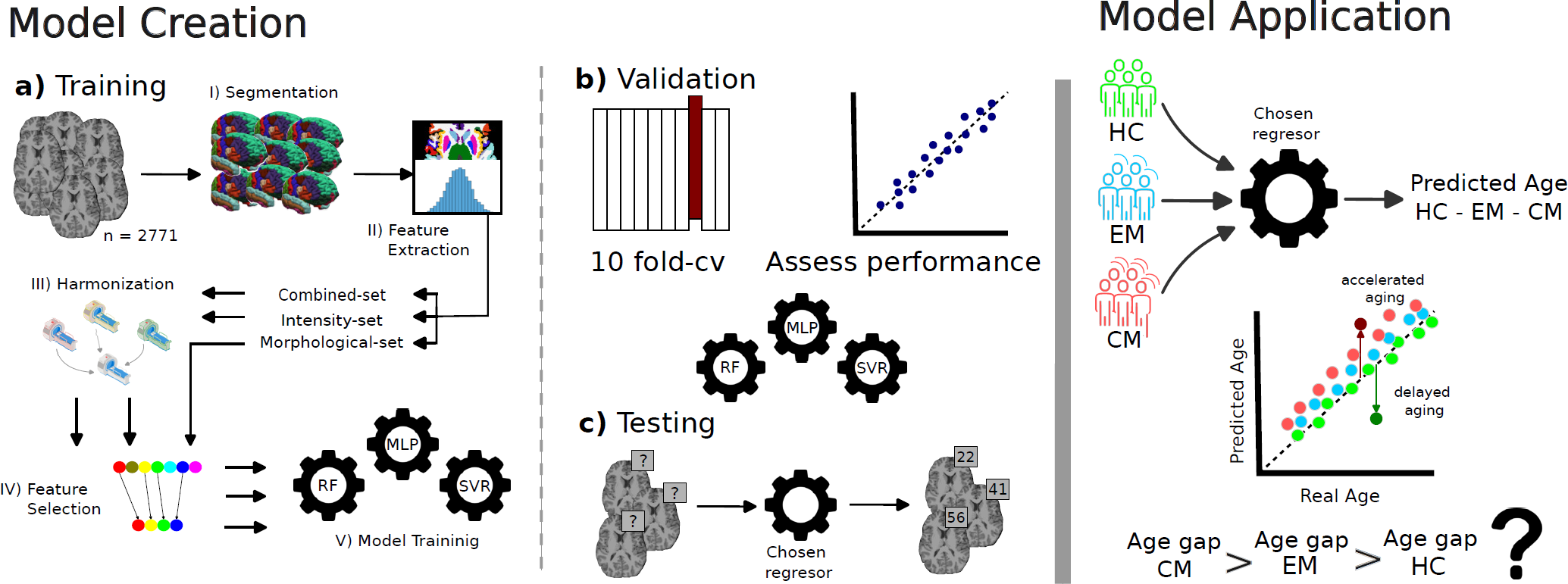
Comprehensive illustration of the methodologies employed for the training of the Brain Age models and the generation of brain predicted-ages. *Model Creation* shows the steps taken to train the Brain Age model on the *Model Creation Dataset* and choose the final model applied on the *Application Dataset* : a) Image processing includes Fastsurfer for brain segmentation and extraction of intensity and morphological features, thus building three feature sets: the *Morphological Feature Set*, *Intensity Feature Set* and the *Combined Feature Set*. For each of these feature sets, a feature selection procedure is performed in a 10-fold cross-validation scheme creating feature sets of 20, 30 and 40 features to feed the machine learning models (SVR, RF and MLP) for each fold. b) Validation is performed to select the best combination of feature set size and machine learning technique. c) Test on the Model *Model Creation Dataset* to assess performance of the Brain Age prediction model. *Model Application* depicts the use of the chosen model on the patient and healthy groups. Brain Age Gap is calculated as the difference between the predicted and the actual age. Differences in Brain Age Gap are then analyzed.

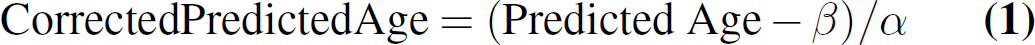

This approach was repeated for each of the aforementioned feature sets. The training procedure was performed using the scikit-learn Python library for machine learning (39). The SVR and RF models were imported from the library while the MLP was implemented using PyTorch (40). Details of the MLP implementation and the hyperparameters for each model are described in Supplementary Table 3.

### Participants

A total of 247 subjects were included in this study, divided into healthy controls (HC, n=82), EM (n=91), and CM patients (n=74). This dataset, on which the Brain Age model previously described was applied, will be hereinafter referred to as *Application Dataset*. Patients were recruited from the outpatient headache unit at the Hospital Clínico Universitario de Valladolid (Spain), a public tertiary care institution that accepts patients from both secondary care and primary care. Inclusion criteria were: a) migraine diagnosis using the third edition of the International Classification of Headache Disorders (ICHD-3) beta and ICHD-3 criteria (1, 41); b) a stable clinical state in the last six months; and c) expressed willingness to partake in the study, coupled with the voluntary signing of the informed consent document. We excluded patients with the following conditions: a) high-frequency episodic migraine, with 10 to 14 headache days per month; b) other painful conditions; c) known major psychiatric diseases (described as anamnesis or the presence of depression or anxiety in the Hospital Anxiety and Depression Scale (42)); d) other neurological diseases; e) drug or substance abuse; and f) pregnancy. At the time of inclusion, no preventive treatment was given to the patients. Participants were requested to complete a headache diary and were diagnosed with EM if they experienced 10 headache days per month or less and CM if they met the ICHD-3 criteria.

Age-and sex-matched HC were recruited through hospital and university colleagues, as well as ads at these institutions, using convenience sampling and snowball sampling. No HC were included if they had a current or previous history of migraine, or if they had any other neurological or mental disorder following the same exclusion criteria as for migraine patients.

We gathered sociodemographic and clinical data from all patients, including migraine illness duration (years) and headache and migraine frequency (days per month).

The study was approved by Hospital Clínico Universitario de Valladolid’s local Ethics Committee (PI: 14-197). All participants read and signed a written consent form before their participation.

### Image acquisition and processing

High-resolution 3D T1w MRI data were acquired for all subjects using a Philips Achieva 3T MRI unit (Philips Healthcare, Best, the Netherlands) with a 32-channel head coil in the MRI facility at the Universidad de Valladolid (Spain). Acquisition parameters were the following: Turbo Field Echo (TFE) sequence, repetition time (TR) = 8.1 ms, echo time (TE) = 3.7 ms, flip angle=8*o*, 256 *×* 256 matrix size, 1 *×* 1 *×* 1 mm^3^ of spatial resolution, and 160 sagittal slices covering the whole brain. Image acquisitions for migraine patients were performed during interictal periods (defined as at least 24 hours from the last migraine attack). Details about the acquisition protocols of each public dataset are described in Supplementary Table 4. If more information is required, further details can be found in each portal of the databases used.

Following the image acquisition, image segmentation, feature extraction and harmonization were also performed on the *Application Dataset* as described for the creation of the Brain Age model. Next, Brain Age was estimated for each participant, including correction from the regression dilution. Since we conducted a 10-fold cross-validation for the training, validation and testing of the Brain Age model, an ensemble formed with the average result of the trained model from each fold was used to obtain the final prediction. Finally, the Brain Age Gap was calculated as the difference between the corrected predicted age and the chronological age of each in-dividual.

### Model interpretation

The significance of each imaging feature in the Brain Age estimation was evaluated using SHapley Additive exPlanations (SHAP) (43). SHAP is a game-theory-based model-agnostic explanation method for machine learning models that evaluates the contribution of each feature to a given prediction. By employing this approach, a group-level comparison of distinct brain imaging features can be conducted to determine their significant contribution to age prediction. Additionally, the evaluation of the influence of individual features on each participant’s Brain Age prediction is made possible, as exemplified in the study conducted by Ballester et al. (44).

The SHAP value for a particular feature for a specific prediction can be interpreted as the difference in the prediction when that feature is omitted from the model. SHAP values reinterpret complex models as a linear function:

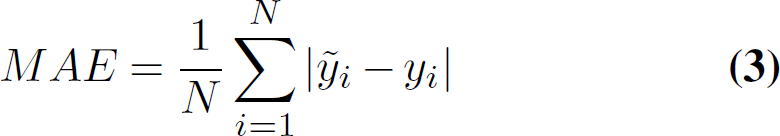

where *z’* is a simplified version of the input features of the model, *φ*_0_ is a reference value of the model (in our case is a value close to the average age of the training data), and *φ_i_*, the attribute effect of the feature which deviates the prediction from the reference value. In a database with N participants and M features, for example, SHAP generates an *N × M* matrix, where each value represents the contribution of feature *m* to the prediction of participant *n*.

We calculated the SHAP value for each subject for a deeper understanding of the regressors. Since many features are repeated across the different regressors, we summed up the contribution of repeated features into a single value. The final matrix was divided by 10 since our ensemble model is the average of the results of the 10 regressors trained during the 10-fold cross-validation.

Once we had the final matrix, we aggregated the values for each of the groups considered (HC, EM and CM) by summing up the absolute values of the matrix along the participant’s axis. The best 15 features in terms of their absolute contribution for each group were selected for each model of the ensemble. Unique features among the three groups studied were selected as the most informative features.

### Statistical analysis

The performance evaluation of the Brain Age models was conducted using two metrics: the MAE and Pearson’s correlation coefficient (*r*). The MAE was calculated as the average of the absolute values of the residuals, which were obtained by subtracting the predicted age from the actual age for each individual in the group. The MAE serves as a comprehensive measure of the prediction error across the entire group, with lower values indicating a better fit. On the other hand, Pearson’s correlation coefficient measures the strength and direction of the linear relationship between the predicted ages and the real ages. Higher values of *r* indicate a better fit of the model. The specific formulas for these metrics can be found in equations Eq. (3) and Eq. (4). Further exploration of these performance metrics can be found in the work from de Lange et al. (38).

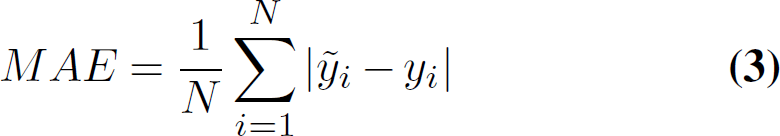

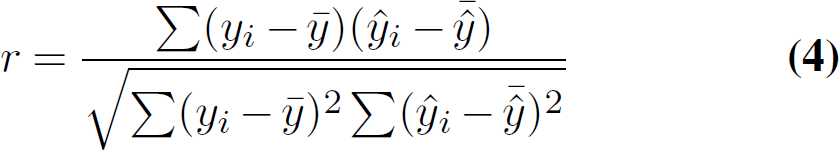

We assessed the normality and homogeneity of variance for age and duration of migraine in the *Application Dataset* using the Kolmogorov-Smirnov test and Levene’s test for equality of variances, respectively. If the null hypothesis was not rejected in both tests, we performed a one-way analysis of variance (ANOVA) to determine significant differences in the ages of the three groups. Gender-significant differences were identified using a chi-square test. For comparing clinical characteristics between migraine patients (i.e., duration of migraine history in years for both groups of patients), we used a two-tailed unpaired t-test if the null hypothesis was not rejected by the Kolmogorov-Smirnov; alternatively, we used the Mann-Whitney U test.

We performed an analysis of covariance (ANCOVA) on the Brain Age Gap results for each pair of groups, adding eTIV and sex as covariates. To verify that the Brain Age Gap calculated for each group was approximately normal and that the variances between groups were comparable, we performed the Kolmogorov-Smirnov test and the Levene test. In the case of a negative Levene’s test, we verified that the variance ratio did not exceed 2 (45). We reset the P value threshold correcting for multiple comparisons using the Bonferroni correction method (P threshold = 0.0167).

Regarding the model interpretation, We conducted a Kruskal-Wallis test on the SHAP values obtained for each of the highly important features of each regressor trained to analyze differences in feature importance among the studied groups. A non-parametric test was chosen due to the non-normality of the SHAP values. To account for multiple comparisons, we applied the Benjamini-Hochberg correction method. We performed pairwise comparisons using the post-hoc Connover-Iman test, correcting its p-values for multiple comparisons using the Benjamini-Hochberg method if the Kruskal-Wallis Test was significant.

Finally, we computed the Pearson’s correlation coefficient between the Brain Age Gap and the clinical characteristics of the migraine groups, correcting for multiple comparisons using the Benjamini-Hochberg method. All statistical tests were conducted in Python.

## Results

### Demographics

There were no significant differences between the groups (HC, EM and CM) in the *Application Dataset* regarding age or sex. Table 2 shows the demographic and clinical characteristics of the dataset, while Figure 2 shows the age distribution of the subjects, together with those in the *Model Creation Dataset*.

**Table 2:** demographic and clinical characteristics for the *Application Dataset*. Not all patients completed the headache diary. Complete data was available from 87 EM patients and 72 CM patients.

|  | HC (n = 82) | EM (n = 91) | CM (n = 74) | Statistical Test |
| --- | --- | --- | --- | --- |
| Woman N°. (%) | 68 (82.9 %) | 75 (82.4 %) | 68 (91.9 %) | sex - $\chi^2 = 3.56$, P = 0.17<sup>†</sup> |
| Age, y | 35.7 ± 12.0 | 36.4 ± 9.9 | 37.8 ± 9.7 | age - ANOVA = 0.86, P = 0.43<sup>‡</sup> |
| **EM n = 87; CM n = 72;** |  |  |  |  |
| Duration of migraine history, y |  | 14.1 ± 10.7 | 19.8 ± 10.7 | t = -3.32, P = 0.001<sup>§</sup> |
| Duration of Chronic migraine, mo |  |  | 28.4 ± 35.3 |  |
| Headache frequency d/mo |  | 3.5 ± 2.1 | 23.5 ± 6.0 | U = 66, P < 0.001 |
| Migraine frequency d/mo |  | 3.6 ± 2.0 | 13.6 ± 6.8 | U = 225.5, P < 0.001 |

Data expressed as mean ± SD
<sup>†</sup>Chi-square test
<sup>‡</sup>ANOVA
<sup>§</sup>Two-tailed, unpaired Student *t* test
<sup>||</sup>Mann-Whitney U test

**Figure 2:**
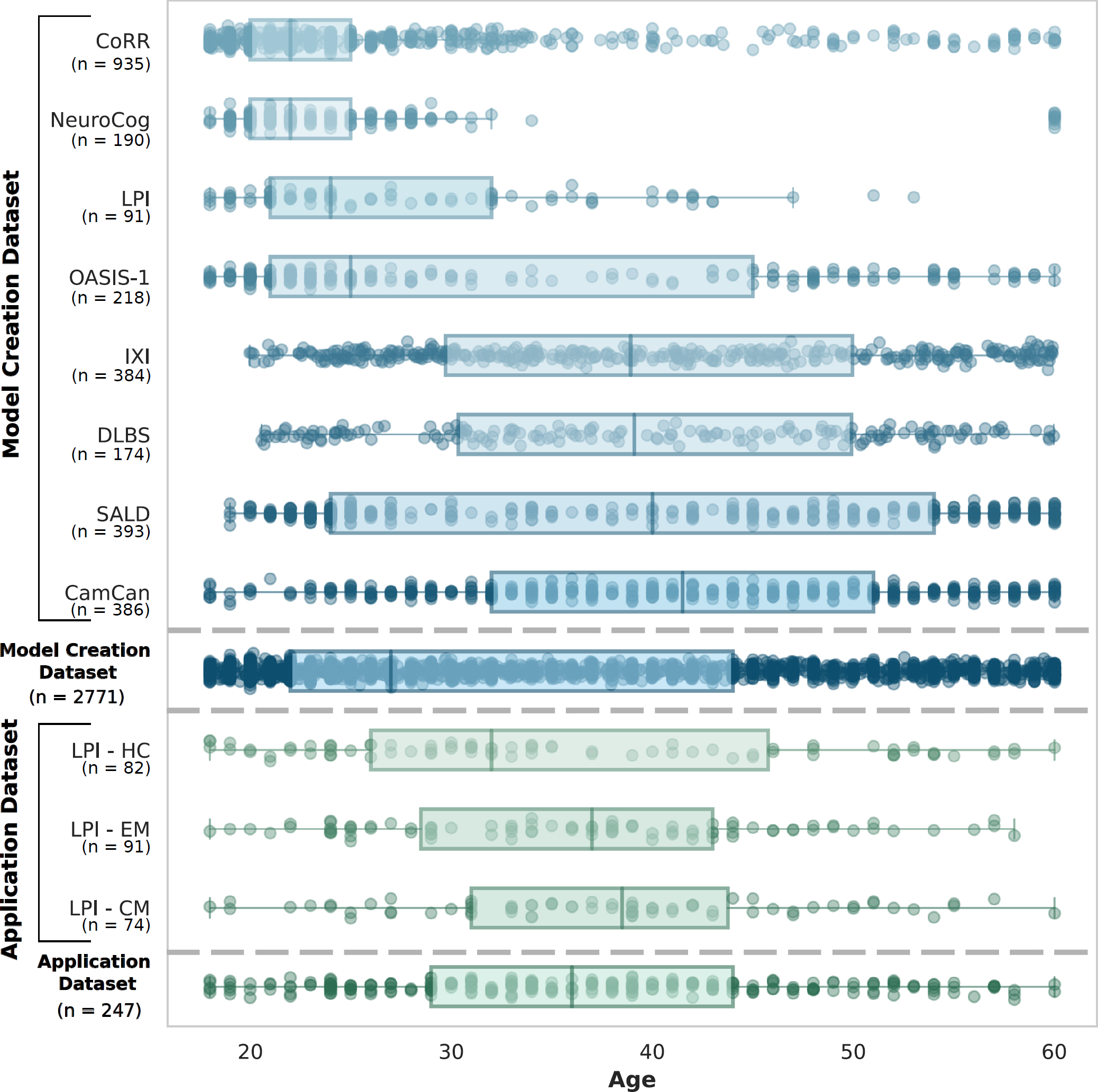
Age distributions of studies in the *Model Creation Dataset* and the *Application Dataset*, ordered by median age.

### Performance of the Brain Age model

Table 3 provides a summary of the validation results for the models tested with the three feature sets studied after age bias correction. Results before the bias correction can be found in Supplementary Table 5. The ensemble model formed by MLPs with a set of 40 selected features provided the greatest performance among the evaluated models in all feature sets and was therefore selected to perform all the Brain Age predictions whose results are described next.

**Table 3:** Validation results for the three regressors tested. Results are given as the average and the standard deviation of the values obtained from each fold of the 10-fold cross-validation scheme after age bias correction. The values in bold show the combination with the best result.

|  |  | SVR |  | RF |  | MLP |  |
| --- | --- | --- | --- | --- | --- | --- | --- |
|  |  | MAE | r | MAE | r | MAE | r |
| Combined Feature Set | 20 features | 7.40 ± 0.50 | 0.82 ± 0.02 | 6.59 ± 0.51 | 0.84 ± 0.02 | 5.99 ± 0.55 | 0.86 ± 0.02 |
|  | 30 features | 7.21 ± 0.50 | 0.83 ± 0.02 | 6.59 ± 0.48 | 0.84 ± 0.02 | 5.81 ± 0.47 | 0.86 ± 0.02 |
|  | 40 features | 7.10 ± 0.42 | 0.83 ± 0.02 | 6.58 ± 0.48 | 0.84 ± 0.02 | **5.79 ± 0.38** | **0.87 ± 0.01** |
| Morphological Feature Set | 20 features | 8.49 ± 0.66 | 0.78 ± 0.02 | 8.75 ± 1.05 | 0.76 ± 0.03 | 7.46 ± 0.85 | 0.80 ± 0.02 |
|  | 30 features | 8.18 ± 0.65 | 0.79 ± 0.02 | 8.62 ± 0.93 | 0.77 ± 0.03 | 7.31 ± 0.76 | 0.81 ± 0.02 |
|  | 40 features | 7.96 ± 0.55 | 0.80 ± 0.02 | 8.50 ± 0.74 | 0.77 ± 0.02 | **7.13 ± 0.52** | **0.81 ± 0.02** |
| Intensity Feature Set | 20 features | 9.25 ± 0.67 | 0.75 ± 0.02 | 9.12 ± 0.68 | 0.75 ± 0.02 | 8.39 ± 0.76 | 0.77 ± 0.02 |
|  | 30 features | 9.15 ± 0.75 | 0.75 ± 0.02 | 9.10 ± 0.75 | 0.76 ± 0.02 | 8.33 ± 0.93 | 0.78 ± 0.03 |
|  | 40 features | 9.09 ± 0.65 | 0.76 ± 0.02 | 9.19 ± 0.77 | 0.75 ± 0.02 | **8.24 ± 0.68** | **0.78 ± 0.02** |

For the test data, training on the *Model Creation Dataset*, the Brain Age model working with the *Combined Feature Set* obtained a MAE and r of 5.95 years and 0.85. On the *Application Dataset*, this same model yielded an MAE and Pearson’s correlation of 6.26 years and 0.84 for the HC group. With regard to the Brain Age model working only with the *Morphological Feature Set*, its performance was MAE = 7.12 years and r = 0.80 on the *Model Creation Dataset*, and MAE = 7.83 years and r = 0.74 on the *Application Dataset* for the HC group. Finally, the Brain Age model working only with the *Intensity Feature Set*, it yielded MAE = 8.19 years and r = 0.78 on the *Model Creation Dataset*, and MAE = 9.27 and years r = 0.64 on the *Application Dataset* for the HC group.

### Brain Age Gap in migraine

Using the *Combined Fea-ture Set*, CM patients exhibited a statistically significant increased Brain Age Gap (average +4.16 vs -0.52 years, P = 0.010) compared to HC. EM patients showed an intermediate Brain Age Gap (average +1.21 years), and neither comparisons with HC nor CM yielded statistical significance. Figure 3 graphically depicts these results, together with scatter-plots showing the brain-predicted age and the chronological age in both the *Model Creation Dataset* and the *Application Dataset*.

**Figure 3:**
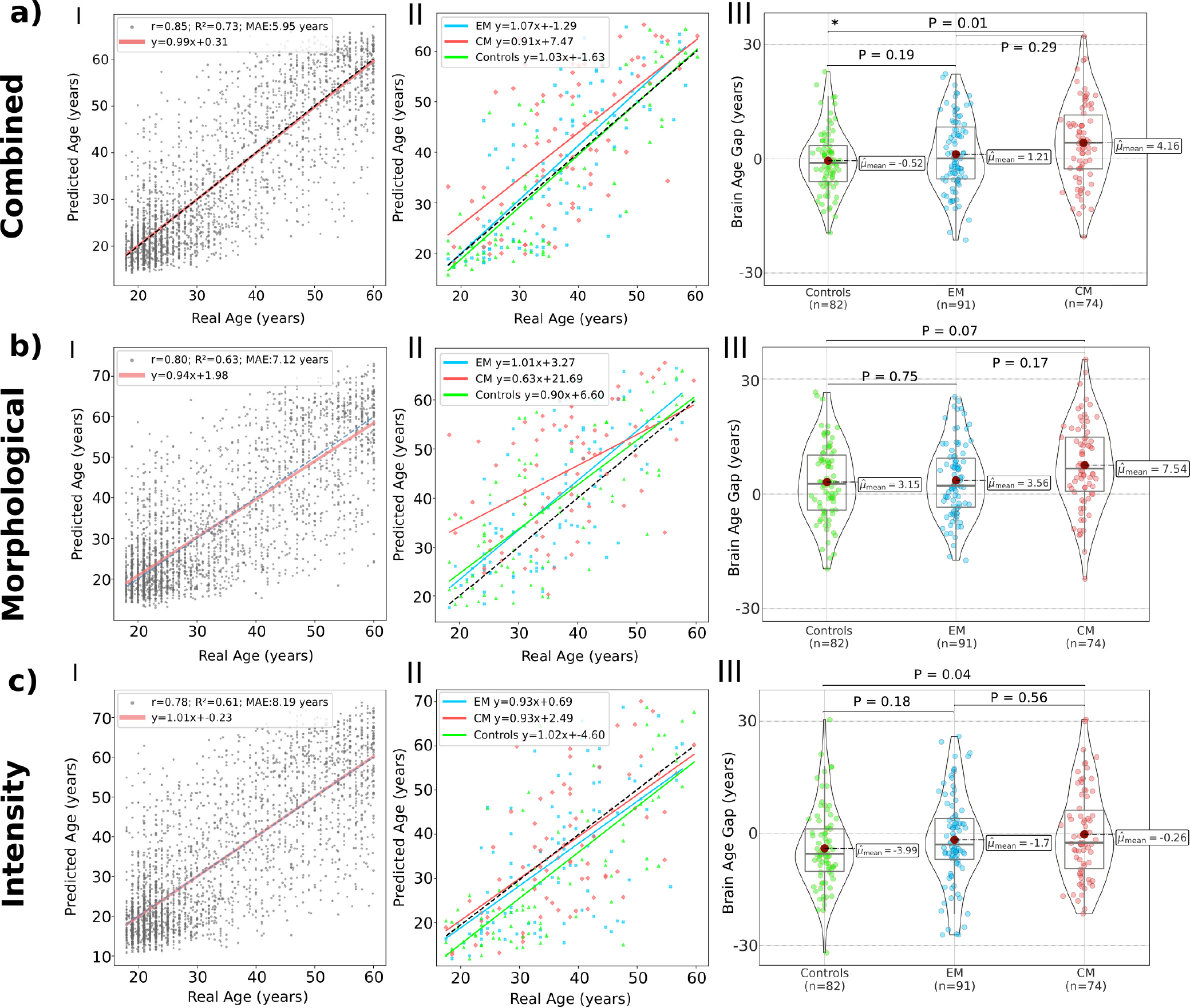
The results of each of the regressors build are shown. The MLPs selecting 40 features demonstrated the best results in validation for all the feature sets. For each trained regressor on every feature set: I) Ensemble MLPs result in the test set of each fold of *Model Creation Dataset*. II) Distribution of the Brain Age Gap values obtained for each of the studied groups. III) Brain Age Gap for the three groups. Statistical significance is denoted by an asterisk (*) to indicate significant findings.

When only employing the *Morphological Feature Set*, CM showed an increased Brain Age Gap with respect to both HC and EM (+7.54 vs 3.15 and 3.56 years, respectively), although these differences were not statistically significant (P = 0.07 and P = 0.17). Interestingly, the average Brain Age Gap for HC and EM were very similar in this case.

Finally, when only employing the *Intensity Feature Set*, CM again showed an increased Brain Age Gap with respect to HC and EM, although differences were not statistically significant in this case either. Figure 3 and Table 4 depict these results.

**Table 4:**
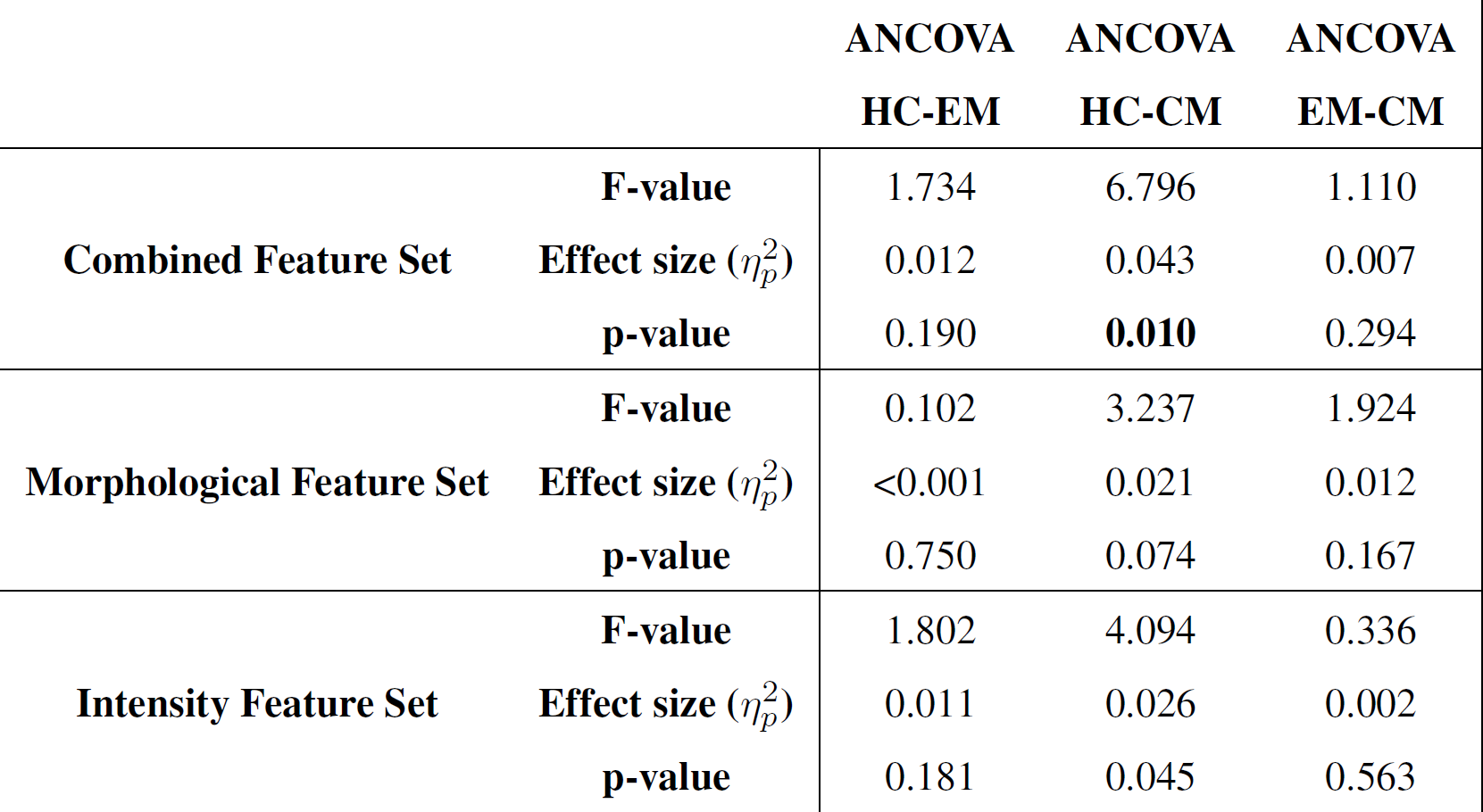
ANCOVA results for Brain Age Gap calculated for each regressor trained. Normality and equality of variances were tested before applying the ANCOVA. Age, sex, and eTIV were included as covariates. The *η*^2^ effect sizes were small (*η*^2^ *<* 0.06) for all comparisons. Statistically significant elements are shown in bold.

|  |  | ANCOVA | ANCOVA | ANCOVA |
| --- | --- | --- | --- | --- |
|  |  | HC-EM | HC-CM | EM-CM |
| **Combined Feature Set** | **F-value** | 1.734 | 6.796 | 1.110 |
|  | **Effect size ($\eta_p^2$)** | 0.012 | 0.043 | 0.007 |
|  | **p-value** | 0.190 | **0.010** | 0.294 |
| **Morphological Feature Set** | **F-value** | 0.102 | 3.237 | 1.924 |
|  | **Effect size ($\eta_p^2$)** | <0.001 | 0.021 | 0.012 |
|  | **p-value** | 0.750 | 0.074 | 0.167 |
| **Intensity Feature Set** | **F-value** | 1.802 | 4.094 | 0.336 |
|  | **Effect size ($\eta_p^2$)** | 0.011 | 0.026 | 0.002 |
|  | **p-value** | 0.181 | 0.045 | 0.563 |

### Model interpretation

Following the SHAP procedure described in Methods, 16 features from the regressor trained on the *Combined Feature Set* were selected. Among them, SHAP values differed significantly between CM patients and HC for the left hemisphere lateral orbitofrontal cortex gray matter volume, left hemisphere superior frontal gyrus gray matter volume and the left hemisphere Insula average thickness (*p <* 0.001 for all cases). For the first two features, the left hemisphere lateral orbitofrontal cortex gray matter volume and the left hemisphere superior frontal gyrus gray matter volume, significant differences were also found in the SHAP values between EM and CM (*p <* 0.01 in both cases). No other significant differences were found for the remaining characteristics among the studied groups. These results are graphically depicted in Figure 4.

**Figure 4:**
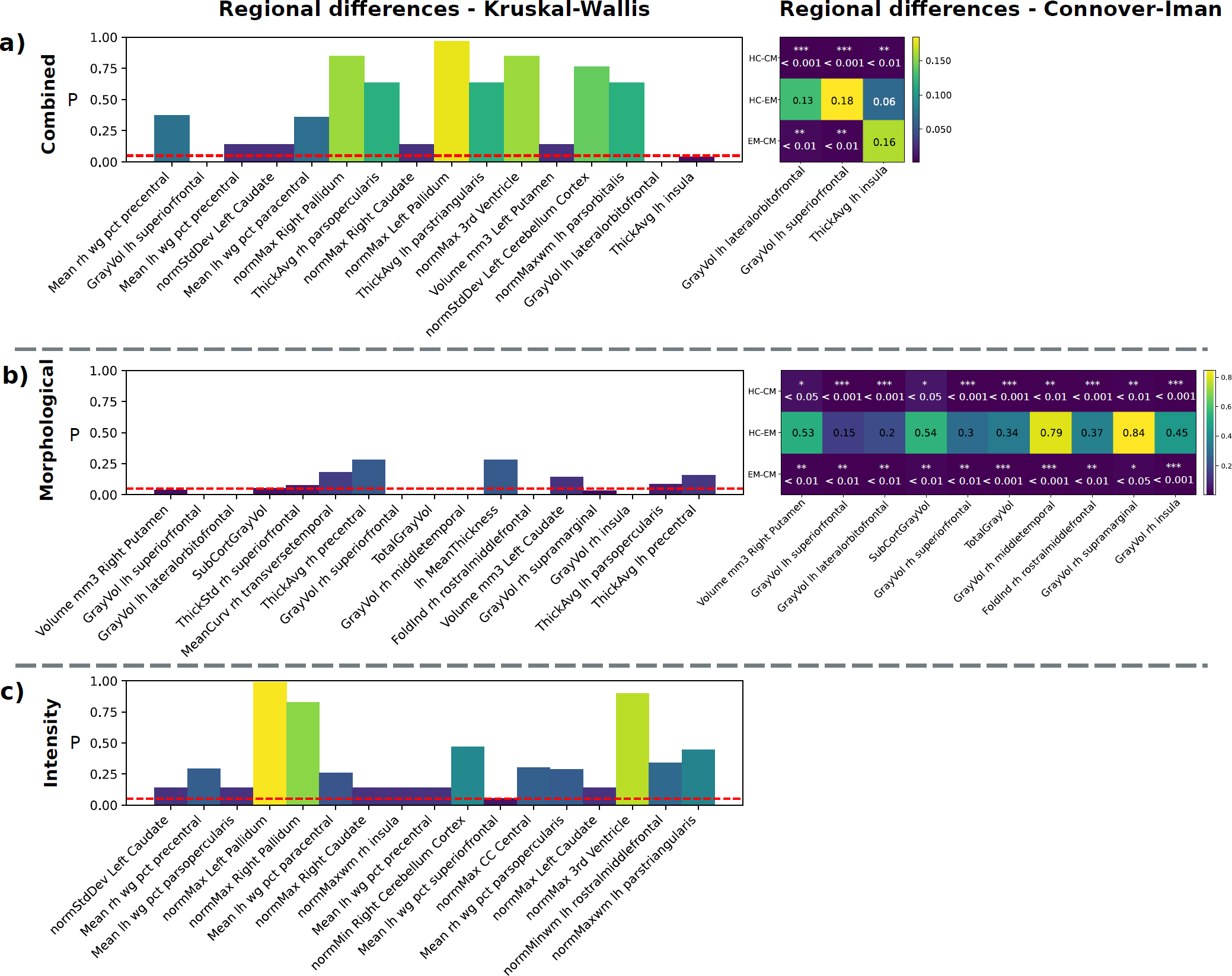
P-values derived from the Kruskal-Wallis test and the post-hoc Connover-Iman test for each of the most significant characteristics chosen for each regressor. Features are ranked from highest importance in the HC group to lower, left-right. a) A total of 16 unique features were selected for the combined regressor, from which 3 demonstrated significant differences in the pairwise comparisons. b) A total of 17 unique features were chosen for the regressor trained on the *Morphological Feature Set*. Up to 10 features demonstrated significant differences in importance between groups. c) shows the 17 most significant characteristics for the regressor trained on the *Intensity Feature Set*. No significant differences were found during the Kruskal-Wallis test.

Regarding the regressor trained on the *Morphological Fea-ture Set*, 17 features were selected. Among them, nine features were significantly different between HC and CM, and ten, (the previous one plus one more) were significantly different for the EM-CM comparison. These are: the volume of the right Putamen (HC-CM *p <* 0.05, EM-CM *p <* 0.01), the volume of the superior frontal gyrus of the left hemisphere (HC-CM *p <* 0.001, EM-CM *p <* 0.01), the volume of the lateral orbitofrontal cortex of the left hemisphere (HC-CM *p <* 0.001, EM-CM <0.01), the volume of the subcortical gray matter (EM-CM *p <* 0.01), the volume of the superior frontal gyrus of the right hemisphere (HC-CM *p <* 0.001, EM-CM *p <* 0.01), the total gray matter volume (HC-CM *p <* 0.001, EM-CM *p <* 0.001), the volume of the middle temporal gyrus of the right hemisphere (HC-CM *p <* 0.01, EM-CM *p <* 0.001), the folding index of the rostral middle frontal gyrus of the right hemisphere (HC-CM *p <* 0.001, EM-CM *p <* 0.01), the volume of the supramarginal gyrus of the right hemisphere (HC-CM *p <* 0.01, EM-CM *p <* 0.05) and the volume of the insula of the right hemisphere (HC-CM *p <* 0.001, EM-CM *p <* 0.001). These features and P values are also shown in Figure 4.

Finally, no features from the regressor trained on the *Intensity Feature Set* (among the 17 that were selected) showed significant differences after the Kruskal-Wallis test. Feature importance for each ensemble studied are depicted Supplementary Figure 2, Supplementary Figure 3 and Supplementary Figure 4.

### Relation between Brain Age Gap and clinical charac-teristics

Employing the regressor trained on the *Combined Feature Set*, and both considering EM and CM separately or together, we found no significant association between the Brain Age Gap and headache frequency (CM -*p* = 0.89, ME -*p* = 0.72, both -*p* = 0.30), migraine frequency (CM -*p* = 0.71, EM -*p* = 0.62, both -*p* = 0.62), migraine duration in years (CM -*p* = 0.52, ME -*p* = 0.52, both -*p* = 0.52) or chronic migraine duration (*p* = 0.32). No significant associations were found either when considering the regressors trained on the *Morphological Feature Set* or the *Intensity Feature Set*. Results are shown in Supplementary Figure 5, Supplementary Figure 6 and Supplementary Figure 7.

## Discussion

In this study, we explored for the first time the Brain Age paradigm in migraine patients. To that end, a machine learning model was developed to predict age from brain T1w MRI acquisitions from a large sample of healthy subjects and later applied to a dataset composed of HC, EM and CM. Several important findings emerged from this investigation. First, CM patients showed an increased predicted Brain Age compared to their healthy counterparts. EM patients, on the other hand, appear to have a much milder increase in their predicted Brain Age, which does not reach statistical significance. Second, different behaviors seem to arise when separately considering morphological and intensity-based features. Distinct difference patterns of difference appear when using either morphological or intensity-based features, furthermore, morphological features seem to be key in the statistically significant difference found. Finally, we could not prove an additional effect of headache frequency or duration of migraine disease in patients’ Brain Age. However, careful consideration is necessary for this point. Most patients presented extremely low (less than 5) or high (20 or more) headache frequency values, and some CM patients included in the relatively small sample could transition between EM and CM states annually.

It is well documented that the process of aging induces changes in both the macroscopic and microstructural properties of the brain (46–48). However, much less is known regarding whether and how disease can affect or interact with this process. A few studies have investigated age-related studies in the migraine brain. Chong et al. (10) examined if aging affects cortical thickness differently in patients with migraine compared to age-matched HC, potentially exacerbating cortical thinning in patients with migraine. For EM patients, the study found that patients with migraine experienced age-related thinning in regions that do not thin in HC. This suggests that migraine may be linked to atypical cortical aging. Lisicki et al., on the other hand, (11), employed FGD-PET to investigate possible specific age-related metabolic changes in the brain. They found that for EM patients advancing age was positively correlated to increased metabolism in the brainstem, hippocampus, fusiform gyrus and parahippocampus, regardless of the frequency of migraine or the duration of the disease. Taken together, these results suggest that migraine and aging do interact in the brain, and the nature of these interactions needs to be investigated.

We found a notable difference in the Brain Age Gap between CM patients and HC. The aforementioned studies, however, did not investigate CM patients but rather focused on EM. For these patients, although we identified a trend towards an increased Brain Age Gap with respect to HC, no significant differences were identified.

Our correlation analysis showed no association between the Brain Age Gap and clinical variables such as headache or migraine frequency and duration of the disease. Whereas these types of associations have been found in many conditions within the Brain Age framework (49–51), in other cases no associations were found between the Brain Age Gap and disease severity or duration (52). This is also the case in (53) with migraine patients, although the Brain Age framework was not explicitly employed in that study. Nevertheless, more extensive research including individuals with high-frequency episodic migraine and greater variation in the duration of the migraine may unveil subtle variances that were eluded in our study.

As with other cerebral changes found in migraine, observational studies such as the present one cannot elucidate possible causalities in the increased Brain Age Gap in CM patients. Interestingly, Vidal-Pineiro et al. (53) found no as-sociation between cross-sectional Brain Age and the rate of Brain Age change measured longitudinally. Instead, they found that the Brain Age Gap seems to be related to early-life factors. Although those results were obtained from large datasets composed of healthy subjects, this “stability” feature of the Brain Age Gap for an individual would be consistent with our findings. Rather, an increased Brain Age Gap could be more related to the susceptibility of an individual to suffer from migraine than to the actual disease. Of course, longitudinal studies are needed to elucidate the nature and the evolution of the Brain Age Gap in migraine.

The literature on Brain Age prediction has utilized a variety of methods for creating machine learning models, including feature-based approaches or Deep Learning techniques (12, 54). In any case, accurate models are needed in order to obtain reliable predictions and good sensitivity to disease. We chose to compile a large dataset (*Model Creation Dataset*) consisting of 2,771 MRI scans from different studies and public databases in order to perform training and validation of the model, and to report its accuracy. The accuracy of our model (MAE and Pearson’s correlation coefficient of 5.95 and 0.85, respectively) when tested on the (*Model Creation Dataset*) is comparable to similar approaches in the literature. More importantly, only a slight accuracy reduction was obtained when applying our model to the *Application Dataset* (MAE and Pearson’s correlation coefficient of 6.26 and 0.84, respectively), which indicates a good generalization capability and performance of the feature harmonization method.

Combining different types of features, as we did with the *Combined Feature Set*, is a prevalent technique in computer vision (55–57). The combination of these features is typically accompanied by an enhancement in the performance of the employed machine learning models (58), as is the case with our study in comparison to the performance obtained for the models trained using the *Morphological Feature Set* and the *Intensity Feature Set*. This is typically explained by the lack of correlation between distinct feature spaces. However, the separate results obtained for the Brain Age models trained using the *Morphological Feature Set* and the *Intensity Feature Set* allow to gain more insight into the behavior of CM and EM patients, since they offer complementary viewpoints of the nature of the Brain Age Gap in migraine. When employing the *Morphological Feature Set*, there is virtually no difference between HC and EM, whereas CM shows an increased Brain Age Gap that appears to vanish for older ages (see Figure 3 (b) II). Although further investigation is required to corroborate this, it suggests that morphological changes in the brain that are associated with CM are more prominent at younger ages, but aging then absorbs these alterations. Conversely, the behavior of the Brain Age Gap when using the *Intensity Feature Set* seems more stable across ages (see Figure 3 (c) II).

The interpretation of the Brain Age predicting model through SHAP allows us to better understand which brain imaging features mostly drive the Brain Age prediction, and which are responsible for the differences found. For the regressor trained on the *Combined Feature Set*, we identified a total of 16 features that highly influenced the prediction of the regressor across the studied groups. These characteristics primarily pertain to the frontal cortex (8), which is expected since features related to the frontal and temporal cortices are common in Brain Age models due to their generalized thinning as part of the normal process of aging (48). Features related to the size of the ventricles are also common in Brain Age models, but not so much in our case (two features) since the increase in the volume of the ventricles is more pronounced after the sixth decade of life (47), while the subjects employed in our study were only between 18 and 60 years old. Ten out of the 16 most significant features were intensity-based, likely representing changes in the tissue microstructure (59).

Three of the brain imaging features found to be most relevant for the Brain Age estimation showed significant differences between CM patients and HC for the regressor trained on the *Combined Feature Set*. Additionally, regarding the regressor trained on the *Morphological Feature Set*, significant differences between CM and HC were found in nine additional features (and nine more features in the comparison between EM and HC). Brain regions related to the identified features have been shown to differ morphologically (6), connectivity-wise (60–62), or both between HC and CM patients and between HC and EM patients. These regions are involved in complex cognitive functions such as information integration or working memory (63, 64) and their alteration may be related to the cognitive changes and other alterations associated with migraine (65, 66).

This study, however, presents several limitations that are worth discussing. First, as a cross-sectional observational study, causality cannot be established, and therefore the interpretation of the results must be taken with caution. Longitudinal studies are needed to elucidate whether susceptibility to migraine, the course of the disease or other factors are responsible for the detected Brain Age Gap between CM and HC, and how this gap evolves with time. Second, our Brain Age model only employs T1w MRI, whereas other modalities such as diffusion MRI have shown to be sensitive to migraine (7). However, research has demonstrated that the integration of various modalities can enhance the precision of Brain Age prediction models (67). Nevertheless, the inclusion of new modalities, despite yielding improved outcomes, can augment the complexity of the overall pipeline. This is particularly exacerbated by the challenges posed by the availability of substantial training data from diverse public datasets and the need for harmonization. Thus, we chose to limit ourselves to T1w scans in this work, although future work will need to include additional modalities such as diffusion MRI, as discussed earlier, or T2-weighted images, given their sensitivity to white matter hyperintensities, a well-known migraine characteristic associated with aging (68). Finally, high-frequency EM patients (10 to 14 headaches per month) were excluded from the study. This decision was made to prevent potentially misclassified patients from skewing the results of the analysis.

## Conclusions

In this study, we analyzed migraine using the Brain Age framework, which consists of training a machine learning model to predict age from MRI scans and later applying the resulting model to a cohort of interest. We found that CM patients exhibit an increased Brain Age Gap (i.e., the difference between the predicted age and the chronological age) compared to HC. A milder Brain Age Gap was found for EM patients, although differences did not reach statistical significance.

Further analysis of the Brain Age model indicated that imaging features that have previously been associated with changes in migraine were among the main drivers of the differences in the predicted age. Also, a separate analysis using only morphological or intensity-based features revealed different patterns, which could represent distinct processes within the alterations that are associated with the migraine brain.

In conclusion, the Brain Age paradigm has shown to be a promising viewpoint for the study of migraine, and future work will be needed to corroborate these findings.

## Data Availability

The public datasets used during the current study are available either online or after reasonable request. The institutional dataset used is also available author upon reasonable request to the corresponding author. The code used for modeling and data analysis is accessible at https://github.com/rafaloz/MigraineBA/

https://fcon_1000.projects.nitrc.org/indi/retro/dlbs.html

http://fcon_1000.projects.nitrc.org/indi/CoRR/html/concept.html

https://openneuro.org/datasets/ds003592

https://www.oasis-brains.org/

http://fcon_1000.projects.nitrc.org/indi/retro/sald.html

https://brain-development.org/ixi-dataset/

https://www.cam-can.org/index.php?content=dataset

## Acknowledgements

The authors are profoundly thankful to the owners of the Neurocognitive aging data release, which is available at OpenNeuro under the identifier “OpenNeuro Dataset ds003592.”, the IXI dataset available at https://brain-development.org/ixi-dataset/, the DLBS dataset and the International Neuroimaging Data-Sharing Initiative Group and the participants of the Consortium for Reliability and Reproducibility (CoRR) for sharing their data publicly so this research was made possible.

Data were provided in part by OASIS OASIS-1: Cross-Sectional: Principal Investigators: D. Marcus, R, Buck-ner, J, Csernansky J. Morris; P50 AG05681, P01 AG03991, P01 AG026276, R01 AG021910, P20 MH071616, U24 RR021382;

Data collection and sharing in part for this project was provided by the Cambridge Centre for Ageing and Neuroscience (CamCAN). CamCAN funding was provided by the UK Biotechnology and Biological Sciences Research Council (grant number BB/H008217/1), together with support from the UK Medical Research Council and University of Cambridge, UK.

The SALD data repository was supported by the National Natural Science Foundation of China (31470981; 31571137; 31500885), National Outstanding young people plan, the Program for the Top Young Talents by Chongqing, the Fundamental Research Funds for the Central Universities (SWU1509383,SWU1509451,SWU1609177), Natural Science Foundation of Chongqing (cstc2015jcyjA10106), Fok Ying Tung Education Foundation (151023), General Financial Grant from the China Postdoctoral Science Foundation (2015M572423, 2015M580767), Special Funds from the Chongqing Postdoctoral Science Foundation (Xm2015037, Xm2016044), Key research for Humanities and social sciences of Ministry of Education (14JJD880009).

In addition, we are extremely grateful to all the patients and participants who volunteered to take part in this study.

## Abbreviations

ANCOVA: Analysis of covariance; ANOVA: Analysis of variance; brain-PAD: brain-predicted age difference; CM: Chronic migraine; CoRR: Consortium for Reliability and Reproducibility; DLBS: Dallas Lifespan Brain Study; EM: Episodic migraine; eTIV: Estimated total intracranial volume; FGD-PET: fluorodeoxyglucose positron emission tomography; fMRI: functional Magnetic Resonance Imaging; HC: Healthy controls; ICHD-3: International Classification of Headache Disorders; IXI: Information eXtraction from Images dataset; MAE: Mean absolute error. MLP: Multilayer perception; NeuroCog: Nerocognitive Aging release; RF: Random forest; SALD: Southwest University Adult Lifespan Dataset; SHAP: Shaply additive explanations; SVR: Support vector regressor; TE: Echo time; TFE: Turbo Field Echo; TR: Repetition time; T1w: T1-weighted;

## Availability of data and materials

The public datasets used during the current study are available either online or after reasonable request. The institutional dataset used is also available author upon reasonable request to the corresponding author. The code used for modeling and data analysis is accessible at https://github.com/rafaloz/MigraineBA/.

## Ethics approval and consent to participate

The research was authorized by Hospital Clínico Universitario de Valladolid’s local Ethics Committee (PI: 14-197).

## Authors’ contributions

RNG and RLG were the major contributors to writing the manuscript. RNG, APG, DGA, and ALG collected and analyzed the data. RLG, SAF, RNG, and APG designed and conceptualized the study, analysed the data and revised the manuscript for intellectual content. All authors read and approved the final manuscript.

## Consent for publication

Not applicable.

## Additional Files

### Additional File 1

**Supplementary Material Table 1:** Features extracted by regions.

| Regions | Features | Total |
| --- | --- | --- |
| **Subcortical regions (45)** | Volume; normalized intensity: mean, standard deviation, minimum, maximum and range | 270 |
| **Cortical regions left and right hemispheres (31x2)** | Area, volume, average thickness, thickness standard deviation, mean curvature, gaussian curvature, folding index, curvature index; White matter gray matter contrast: mean, standard deviation, minimum maximum and range | 806 |
| **White matter left and right hemispheres (32x2)** | Volume; normalized intensity: mean, standard deviation, minimum, maximum and range | 384 |
| **Whole brain features** |  | 19 |
| **Total** |  | **1479** |

**Supplementary Material Figure 1:**
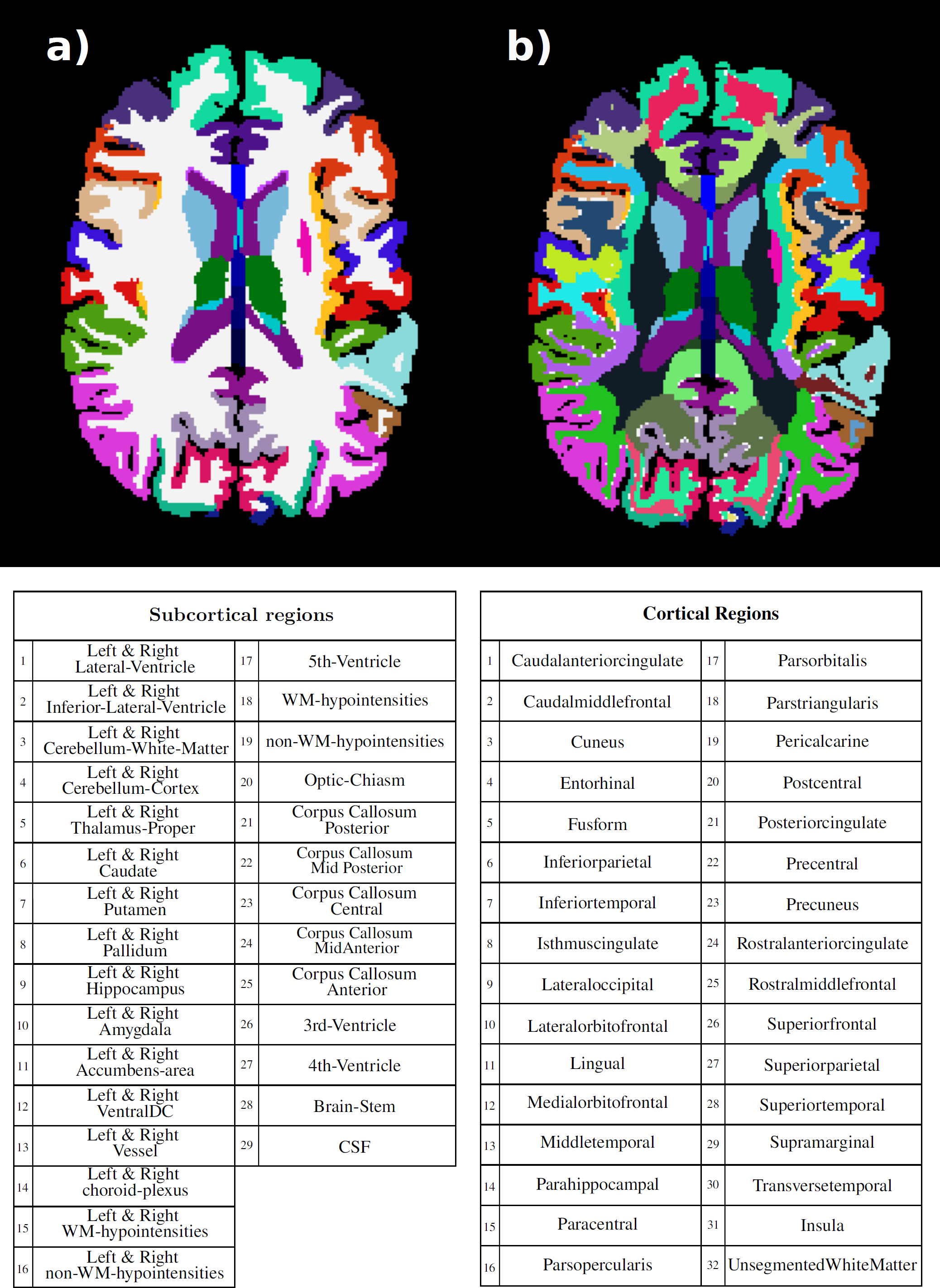
Segmentation example from case 110033 of the CamCAN database. On the left a), the segmentation is shown without the white matter segmentation. Subcortical and cortical regions are divided. On the right b) the segmentation includes white matter segmentation.

**Supplementary Material Table 2:** Features extracted from the whole brain.

| Whole brain features |  |
| --- | --- |
| 1 | Brain Segmentation Volume |
| 2 | Left hemisphere cortical gray matter volume |
| 3 | Right hemisphere cortical gray matter volume |
| 4 | Subcortical gray matter volume |
| 5 | Total gray matter volume |
| 6 | Supratentorial volume |
| 7 | Mask Volume |
| 8 | Number of defect holes in lh surfaces prior to fixing |
| 9 | Number of defect holes in rh surfaces prior to fixing |
| 10 | Estimated Total Intracranial Volume |
| 11 | Left Hemisphere White Surface Total Area |
| 12 | Right Hemisphere White Surface Total Area |
| 13 | Left Hemisphere Cortex Mean Thickness |
| 14 | Right Hemisphere Cortex Mean Thickness |
| 15 | Total cortical gray matter volume |
| 16 | Volume of ventricles and choroid plexus |
| 17 | Left hemisphere cerebral white matter volume |
| 18 | Right hemisphere cerebral white matter volume |
| 19 | Total cerebral white matter volume |

**Supplementary Material Table 3:** Hyperparameters of the regressors trained for the study.

| Regressors | Hyperparameters |
| --- | --- |
| **SVR** | kernel='linear', degree=3, gamma='scale', coef0=0.0, tol=0.001, C=1.0, epsilon=0.1, shrinking=True, cache_size=200 |
| **RF** | n_estimators=100, criterion='squared_error', max_depth=None, bootstrap=True, min_samples_split=2, min_samples_leaf=1, min_weight_fraction_leaf=0.0, max_features=1.0, max_leaf_nodes=None, min_impurity_decrease=0.0, oob_score=False, ccp_alpha=0.0 |
| **MLP** | epochs=500, lr=0.01, weight_decay=0.01, validation_size=0.2, criterion=L1, optimizer=Adam, early_stopping=20 epochs. |

**Supplementary Material Table 4:**
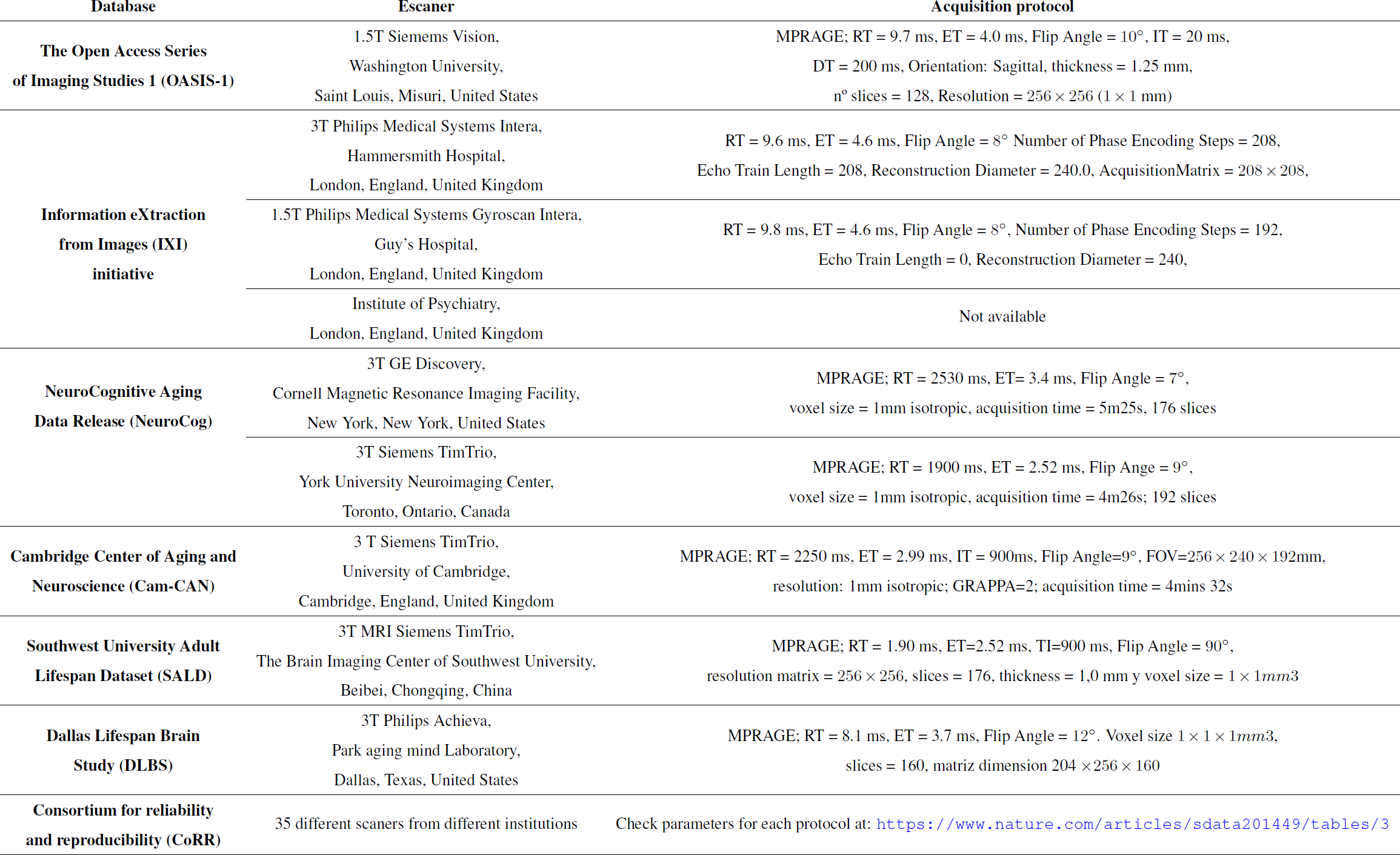
Acquisition parameters for each scanner employed in every database used to construct the Brain Age model.

**Supplementary Material Table 5:** Validation results for the three regressors tested. Results are given as the average and the standard deviation of the values obtained from each fold of the 10-fold cross-validation scheme before age bias correction. The values in bold show the combination with the best result.

|  |  | SVR |  | RF |  | MLP |  |
| --- | --- | --- | --- | --- | --- | --- | --- |
|  |  | MAE | r | MAE | r | MAE | r |
| Combined Feature Set | 20 features | 6.07 ± 0.29 | 0.82 ± 0.02 | 5.51 ± 0.25 | 0.84 ± 0.02 | 5.03 ± 0.29 | 0.86 ± 0.02 |
|  | 30 features | 5.94 ± 0.24 | 0.83 ± 0.02 | 5.54 ± 0.26 | 0.84 ± 0.02 | 4.92 ± 0.25 | 0.86 ± 0.02 |
|  | 40 features | 5.85 ± 0.22 | 0.83 ± 0.02 | 5.55 ± 0.27 | 0.84 ± 0.02 | **4.90 ± 0.21** | **0.87 ± 0.01** |
| Morphological Feature Set | 20 features | 6.68 ± 0.43 | 0.78 ± 0.02 | 6.53 ± 0.54 | 0.76 ± 0.03 | 5.74 ± 0.47 | 0.80 ± 0.02 |
|  | 30 features | 6.52 ± 0.44 | 0.79 ± 0.02 | 6.54 ± 0.46 | 0.77 ± 0.03 | 5.66 ± 0.44 | 0.81 ± 0.02 |
|  | 40 features | 6.37 ± 0.39 | 0.80 ± 0.02 | 6.46 ± 0.42 | 0.77 ± 0.02 | **5.57 ± 0.27** | **0.81 ± 0.02** |
| Intensity Feature Set | 20 features | 6.91 ± 0.41 | 0.75 ± 0.02 | 6.64 ± 0.40 | 0.75 ± 0.02 | 6.17 ± 0.40 | 0.77 ± 0.02 |
|  | 30 features | 6.87 ± 0.46 | 0.75 ± 0.02 | 6.67 ± 0.43 | 0.76 ± 0.02 | 6.13 ± 0.44 | 0.78 ± 0.03 |
|  | 40 features | 6.80 ± 0.38 | 0.76 ± 0.02 | 6.72 ± 0.41 | 0.75 ± 0.02 | **6.06 ± 0.35** | **0.78 ± 0.02** |

**Supplementary Material Figure 2:**
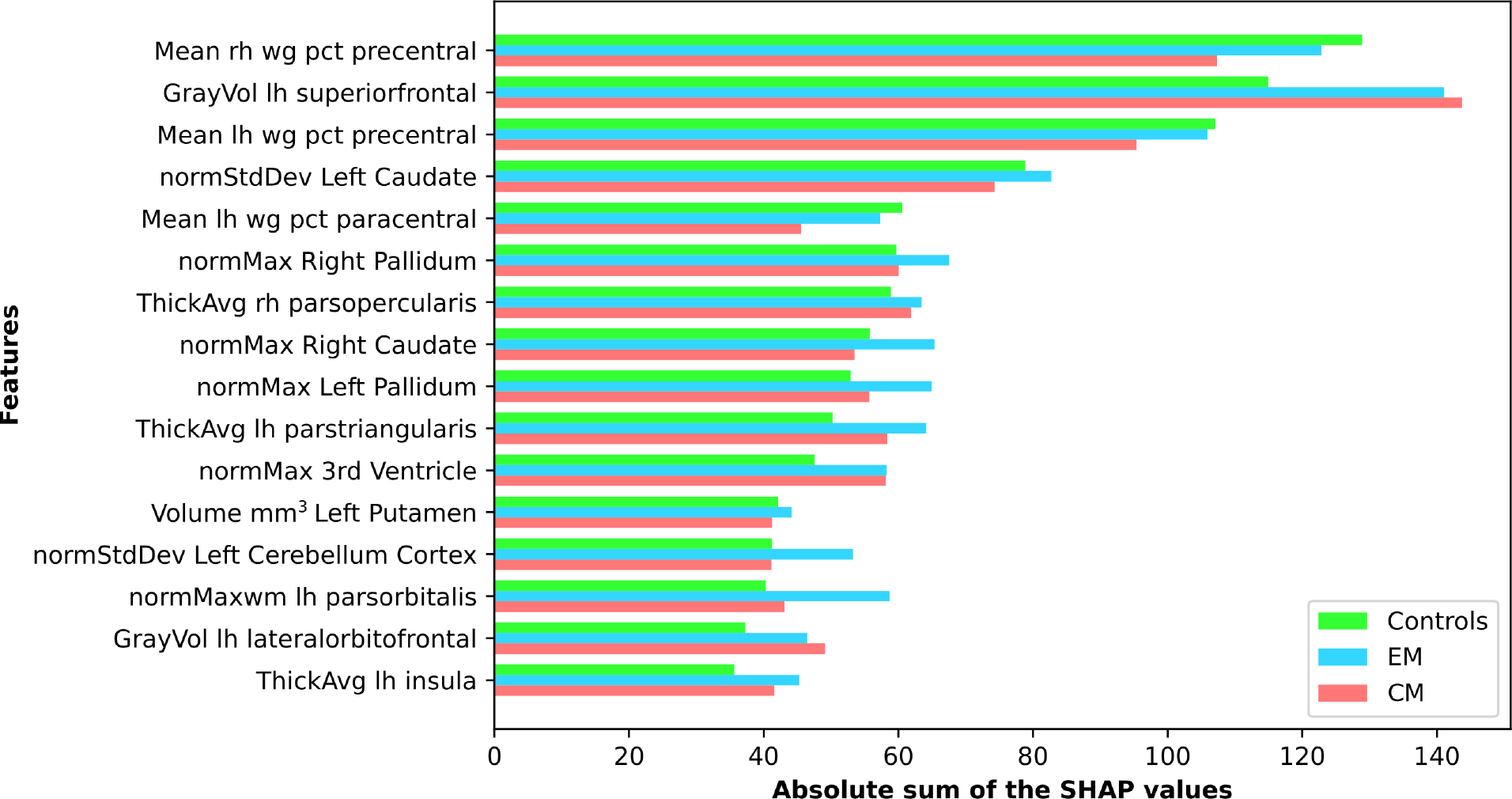
The sum of the absolute SHAP values of each feature for each member of the investigated groups, calculated for the regressor trained on the *Combined Feature Set*. The order of features varies between groups, but the 16 designated features are shared by the groups’ most pertinent features.

**Supplementary Material Figure 3:**
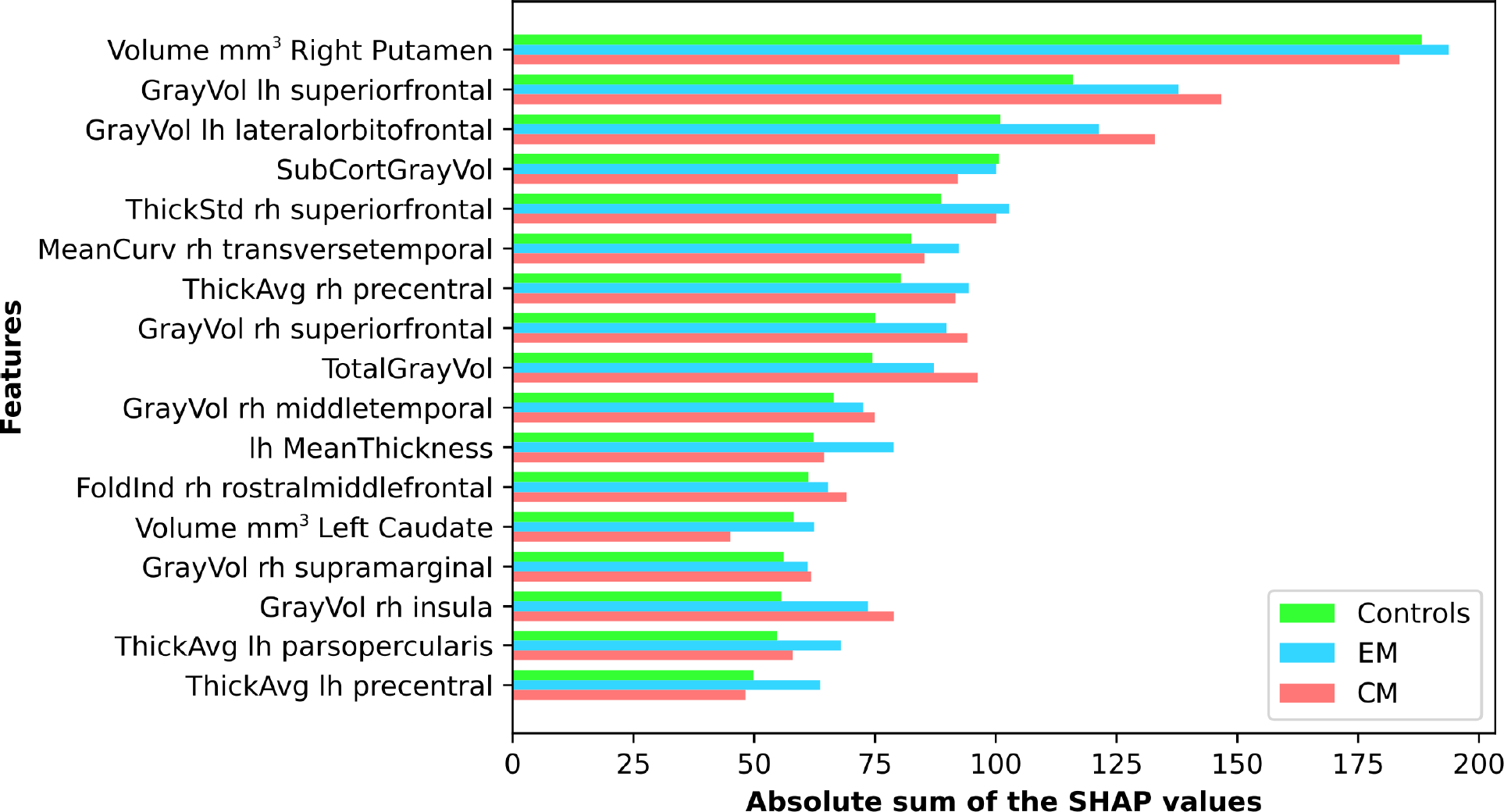
The sum of the absolute SHAP values of each feature for each member of the investigated groups, calculated for the regressor trained on the *Morphological Feature Set*. The order of features varies between groups, but the 17 designated features are shared by the groups’ most pertinent features.

**Supplementary Material Figure 4:**
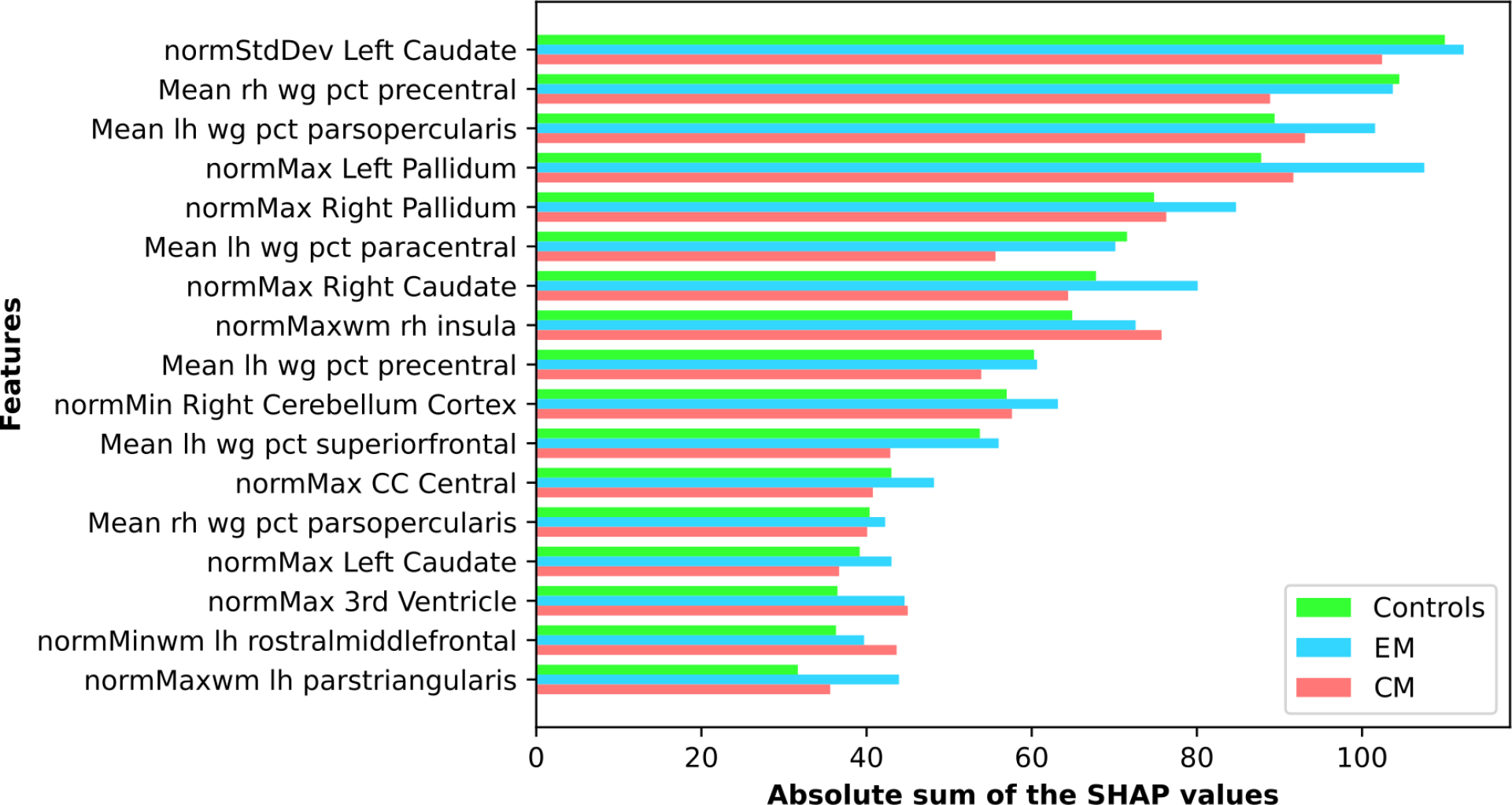
The sum of the absolute SHAP values of each feature for each member of the investigated groups, calculated for the regressor trained on the *Intensity Feature Set*. The order of features varies between groups, but the 17 designated features are shared by the groups’ most pertinent features.

**Supplementary Material Figure 5:**
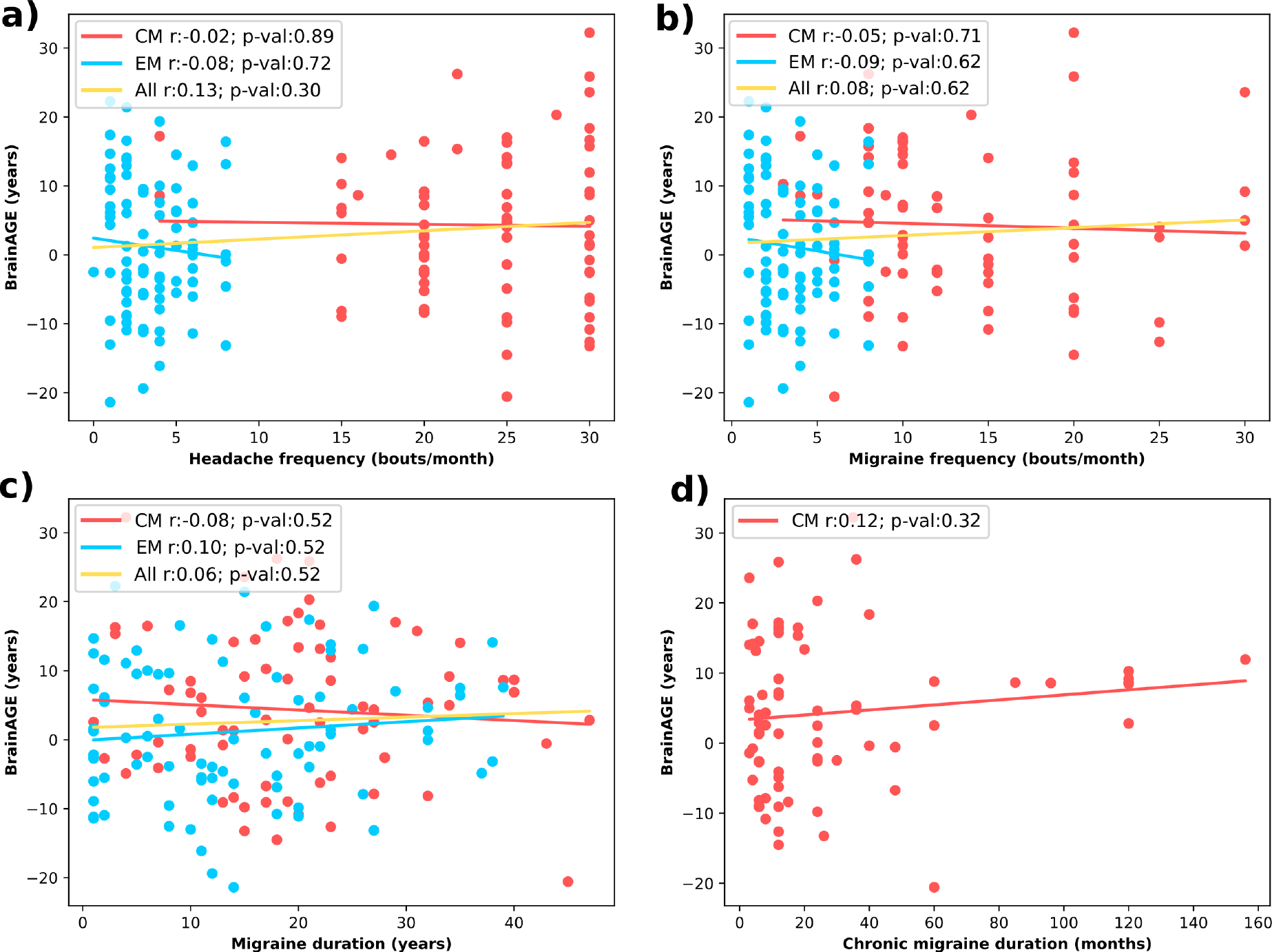
No correlations were found between Brain Age Gap calculated with the regressor trained on the *Combined Feature Set* and the clinical variables studied, a) Brain Age Gap change along with headache frequency, b) Brain Age Gap change along with migraine frequency, c) Brain Age Gap change along with migraine duration, and d) Brain Age Gap change along with chronic migraine duration.

**Supplementary Material Figure 6:**
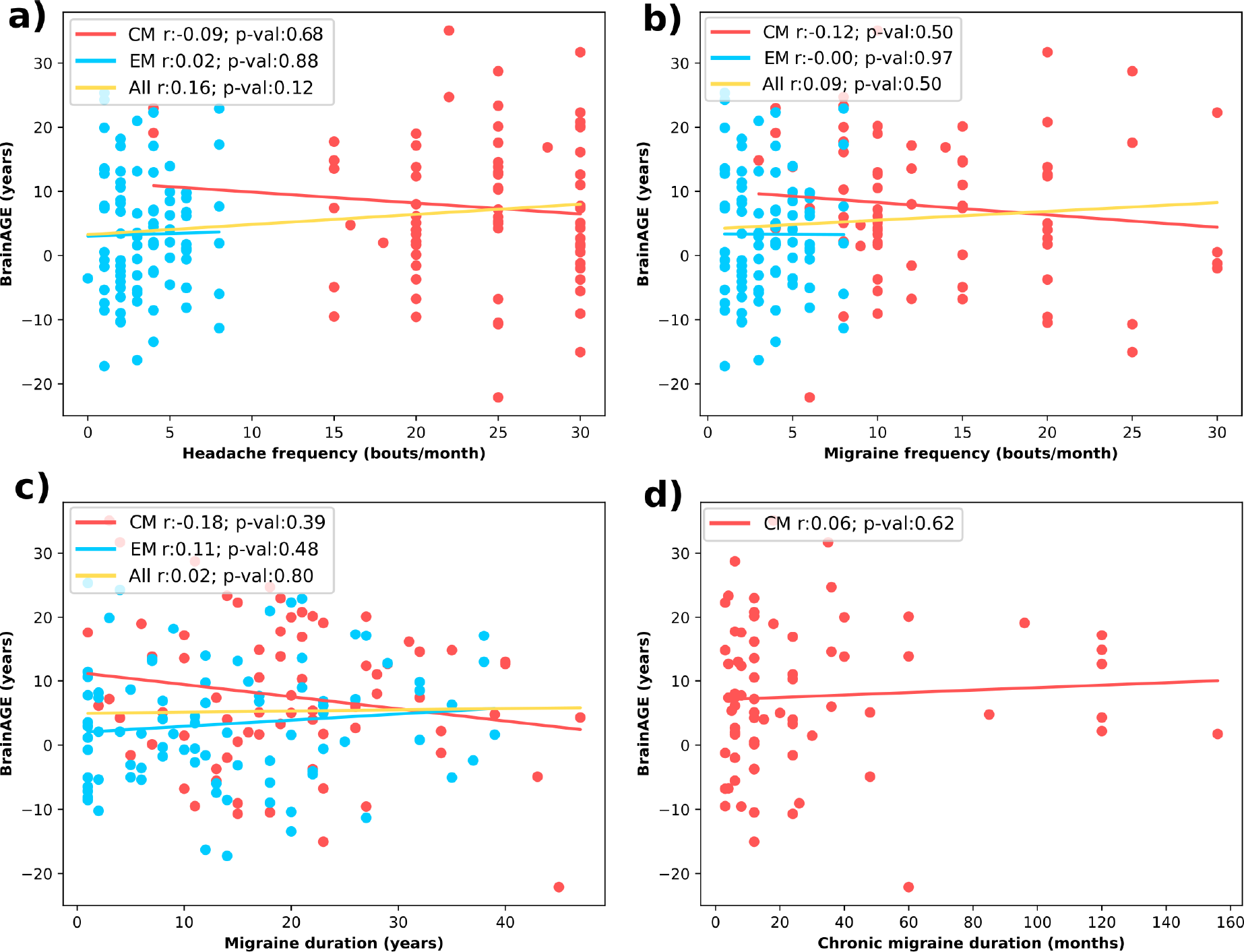
No statistically significant correlation was found between clinical variables and Brain Age Gap when calculated with the regressor trained on the *Morphological Feature Set*, a) Brain Age Gap change along with headache frequency, b) Brain Age Gap change along with migraine frequency, c) Brain Age Gap change along with migraine duration, and d) Brain Age Gap change along with chronic migraine duration.

**Supplementary Material Figure 7:**
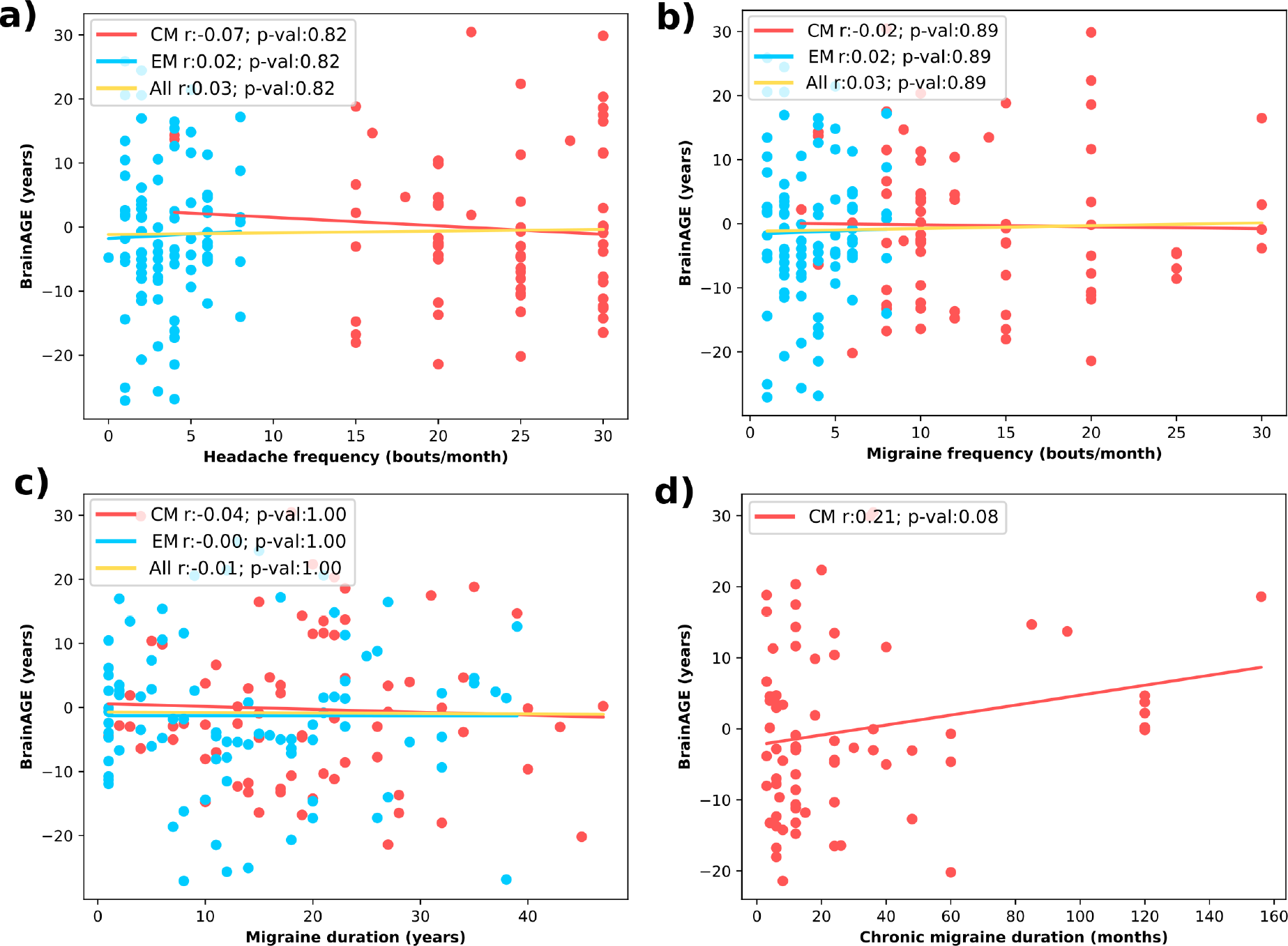
No statistically significant correlation was found between the clinical variables and the Brain Age Gap when calculated with the regressor trained on the *Intensity Feature Set*, a) Brain Age Gap change along with headache frequency, b) Brain Age Gap change along with migraine frequency, c) Brain Age Gap change along with migraine duration, and d) Brain Age Gap change along with chronic migraine duration.

## Graphical Abstract

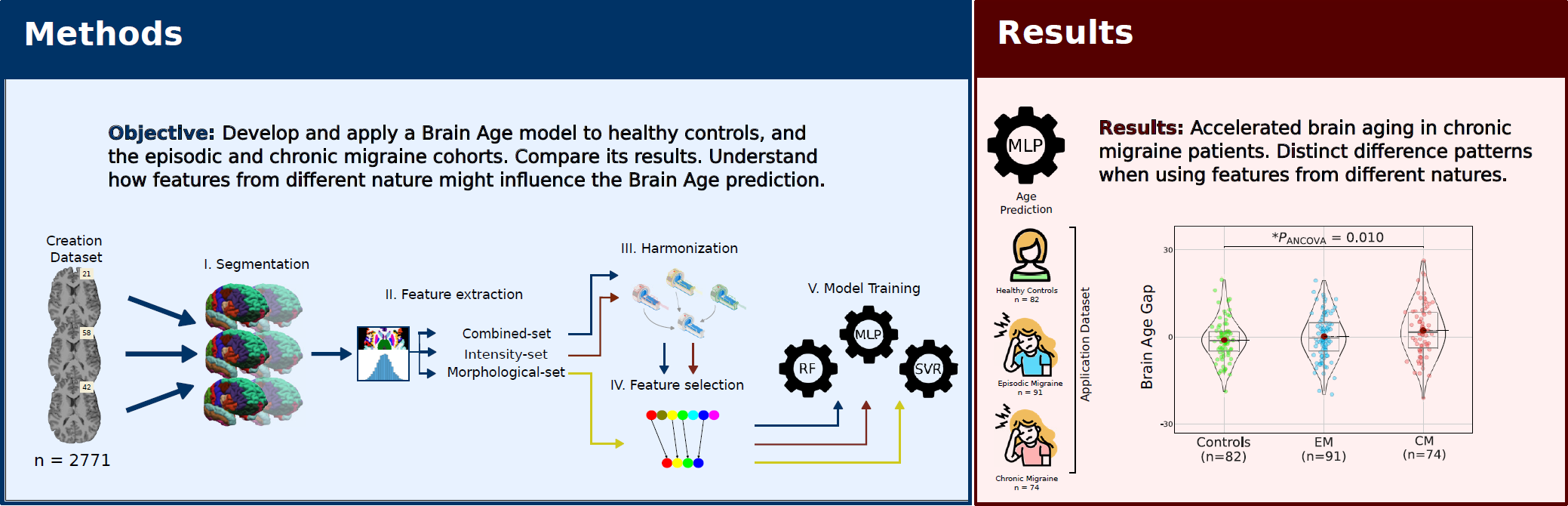

## Bibliography

1. Marcel Arnold. Headache classification committee of the international headache society (IHS) the international classification of headache disorders. Cephalalgia, 38(1):1–211, 2018. doi: 10.1177/0333102417738202.

2. Michel D Ferrari, Peter J Goadsby, Rami Burstein, Tobias Kurth, Cenk Ayata, Andrew Charles, Messoud Ashina, Arn MJM van den Maagdenberg, and David W Dodick. Migraine. Nature reviews Disease primers, 8(1), 2022. doi: 10.1038/s41572-021-00328-4.

3. Z. Jia and S. Yu. Grey matter alterations in migraine: A systematic review and meta-analysis. Neuroimage Clinical, 14:130–140, 2017. doi: 10.1016/j.nicl.2017.01.019.

4. A. Kattem-Husøy, L. Eikenes, A. K. Håberg, K. Hagen, and L. J. Stovner. Diffusion tensor imaging in middle-aged headache sufferers in the general population: a cross-sectional population-based imaging study in the Nord-Trøndelag health study (HUNT-MRI). The Journal of Headache and Pain, 20(1):1–15, 2019. doi: 10.1186/s10194-019-1028-6.

5. T. J. Schwedt, C.-C. Chiang, C. D. Chong, and D. W. Dodick. Functional MRI of migraine. The Lancet Neurology, 14(1):81–91, 2015. doi: 10.1016/S1474-4422(14)70193-0.

6. Álvaro Planchuelo-Gómez, David García-Azorín, Ángel L Guerrero, Margarita Rodríguez, Santiago Aja-Fernández, and Rodrigo de Luis-García. Gray matter structural alterations in chronic and episodic migraine: a morphometric magnetic resonance imaging study. Pain Medicine, 21(11):2997–3011, 2020. doi: 10.1093/pm/pnaa271.

7. Álvaro Planchuelo-Gómez, David García-Azorín, Ángel L Guerrero, Santiago Aja-Fernández, Margarita Rodríguez, and Rodrigo de Luis-García. White matter changes in chronic and episodic migraine: a diffusion tensor imaging study. The Journal of Headache and Pain, 21:1–15, 2020. doi: 10.1186/s10194-019-1071-3.

8. M. J. Lee, B.-y. Park, S. Cho, S. T. Kim, H. Park, and C.-S. Chung. Increased connectivity of pain matrix in chronic migraine: a resting-state functional MRI study. The Journal of Headache and Pain, 20(1):1–10, 2019. doi: 10.1186/s10194-019-0986-z.

9. Tiffany Bell, Akashroop Khaira, Mehak Stokoe, Megan Webb, Melanie Noel, Farnaz Amoozegar, and Ashley D Harris. Age-related differences in resting state functional connectivity in pediatric migraine. The Journal of Headache and Pain, 22(1):1–12, 2021. doi: 10.1186/s10194-021-01274-y.

10. Catherine D Chong, David W Dodick, Bradley L Schlaggar, and Todd J Schwedt. Atypical age-related cortical thinning in episodic migraine. Cephalalgia, 34(14):1115–1124, 2014. doi: 10.1177/0333102414531157.

11. Marco Lisicki, Kevin D’Ostilio, Gianluca Coppola, Vincenzo Parisi, Alain Maertens de Noordhout, Delphine Magis, Jean Schoenen, Felix Scholtes, and Jan Versijpt. Age related metabolic modifications in the migraine brain. Cephalalgia, 39(8):978–987, 2019. doi: 10.1177/0333102419828984.

12. Katja Franke, Gabriel Ziegler, Stefan Klöppel, Christian Gaser, Alzheimer’s Disease Neuroimaging Initiative, et al. Estimating the age of healthy subjects from T1-weighted MRI scans using kernel methods: exploring the influence of various parameters. Neuroimage, 50(3):883–892, 2010. doi: 10.1016/j.neuroimage.2010.01.005.

13. Iman Beheshti, Shiwangi Mishra, Daichi Sone, Pritee Khanna, and Hiroshi Matsuda. T1-weighted MRI-driven brain age estimation in Alzheimer’s disease and Parkinson’s disease. Aging and disease, 11(3):618, 2020. doi: 10.14336/AD.2019.0617.

14. Pedro L Ballester, Maria T Romano, Taiane de Azevedo Cardoso, Stefanie Hassel, Stephen C Strother, Sidney H Kennedy, and Benicio N Frey. Brain age in mood and psychotic disorders: a systematic review and meta-analysis. Acta Psychiatrica Scandinavica, 145(1):42–55, 2022. doi: 10.1111/acps.13371.

15. Katja Franke, Eileen Luders, Arne May, Marko Wilke, and Christian Gaser. Brain maturation: predicting individual BrainAGE in children and adolescents using structural MRI. Neuroimage, 63(3):1305–1312, 2012. doi: 10.1016/j.neuroimage.2012.08.001.

16. Lars Rogenmoser, Julius Kernbach, Gottfried Schlaug, and Christian Gaser. Keeping brains young with making music. Brain Structure and Function, 223(1):297–305, 2018. doi: 10.1007/s00429-017-1491-2.

17. Jason Steffener, Christian Habeck, Deirdre O’Shea, Qolamreza Razlighi, Louis Bherer, and Yaakov Stern. Differences between chronological and brain age are related to education and self-reported physical activity. Neurobiology of aging, 40:138–144, 2016. doi: 10.1016/j.neurobiolaging.2016.01.014.

18. Eileen Luders, Nicolas Cherbuin, and Christian Gaser. Estimating brain age using high-resolution pattern recognition: Younger brains in long-term meditation practitioners. Neuroimage, 134:508–513, 2016. doi: 10.1016/j.neuroimage.2016.04.007.

19. James H Cole, Stuart J Ritchie, Mark E Bastin, Valdés Hernández, S Muñoz Maniega, Natalie Royle, Janie Corley, Alison Pattie, Sarah E Harris, Qian Zhang, et al. Brain age predicts mortality. Molecular psychiatry, 23(5):1385–1392, 2018. doi: 10.1038/mp.2017.62.

20. Y. Cruz-Almeida, R. B. Fillingim, J. L. Riley III, A. J. Woods, E. Porges, R. Cohen, and J. Cole. Chronic pain is associated with a brain aging biomarker in community-dwelling older adults. Pain, 160(5):1119, 2019. doi: 10.1097/j.pain.0000000000001491.

21. P. Sörös and C. Bantel. Chronic noncancer pain is not associated with accelerated brain aging as assessed by structural magnetic resonance imaging in patients treated in specialized outpatient clinics. Pain, 161(3):641–650, 2020. doi: 10.1097/j.pain.0000000000001756.

22. A. J. Johnson, J. Cole, R. B. Fillingim, and Y. Cruz-Almeida. Persistent Non-pharmacological Pain Management and Brain-Predicted Age Differences in Middle-Aged and Older Adults With Chronic Knee Pain. Frontiers in Pain Research, 3:868546, 2022. doi: 10.3389/fpain.2022.868546.

23. Z. Y. Gary, Karim Ly, M., H. T. N. Muppidi, H. J. Aizenstein, and J. W. Ibinson. Accelerated brain aging in chronic low back pain. Brain research, 1755:147263, 2022. doi: 10.1016/j.brainres.2020.147263.

24. The Aging Mind Lab, University of Texas. Dallas lifespan Brain Study, 2023.

25. Xi-Nian Zuo, Jeffrey S Anderson, Pierre Bellec, Rasmus M Birn, Bharat B Biswal, Janusch Blautzik, John Breitner, Randy L Buckner, Vince D Calhoun, F Xavier Castellanos, et al. An open science resource for establishing reliability and reproducibility in functional connectomics. Scientific data, 1(1):1–13, 2014. doi: 10.1038/sdata.2014.49.

26. R Nathan Spreng, Roni Setton, Udi Alter, Benjamin N Cassidy, Bri Darboh, Elizabeth DuPre, Karin Kantarovich, Amber W Lockrow, Laetitia Mwilambwe-Tshilobo, Wen-Ming Luh, et al. Neurocognitive aging data release with behavioral, structural and multi-echo functional MRI measures. Scientific Data, 9(1):1–11, 2022. doi: 10.1038/s41597-022-01231-7.

27. Daniel S Marcus, Tracy H Wang, Jamie Parker, John G Csernansky, John C Morris, and Randy L Buckner. Open Access Series of Imaging Studies (OASIS): cross-sectional MRI data in young, middle aged, nondemented, and demented older adults. Journal of cognitive neuroscience, 19(9):1498–1507, 2007. doi: 10.1162/jocn.2007.19.9.1498.

28. Dongtao Wei, Kaixiang Zhuang, Lei Ai, Qunlin Chen, Wenjing Yang, Wei Liu, Kangcheng Wang, Jiangzhou Sun, and Jiang Qiu. Structural and functional brain scans from the cross-sectional Southwest University adult lifespan dataset. Scientific data, 5(1):1–10, 2018. doi: 10.1038/sdata.2018.134.

29. Biomedical Image Analysis Group, Imperial College London. IXI dataset portal, 2023.

30. Jason R Taylor, Nitin Williams, Rhodri Cusack, Tibor Auer, Meredith A Shafto, Marie Dixon, Lorraine K Tyler, Richard N Henson, et al. The Cambridge Centre for Ageing and Neuroscience (Cam-CAN) data repository: Structural and functional MRI, MEG, and cognitive data from a cross-sectional adult lifespan sample. neuroimage, 144:262–269, 2017. doi: 10.1016/j.neuroimage.2015.09.018.

31. Meredith A Shafto, Lorraine K Tyler, Marie Dixon, Jason R Taylor, James B Rowe, Rhodri Cusack, Andrew J Calder, William D Marslen-Wilson, John Duncan, Tim Dalgleish, et al. The Cambridge Centre for Ageing and Neuroscience (Cam-CAN) study protocol: a cross-sectional, lifespan, multidisciplinary examination of healthy cognitive ageing. BMC neurology, 14(1):1–25, 2014. doi: 10.1186/s12883-014-0204-1.

32. Leonie Henschel, Sailesh Conjeti, Santiago Estrada, Kersten Diers, Bruce Fischl, and Mar-tin Reuter. Fastsurfer-a fast and accurate deep learning based neuroimaging pipeline. NeuroImage, 219:117012, 2020. doi: 10.1016/j.neuroimage.2020.117012.

33. A Klein and J Tourville. 101 labeled brain images and a consistent human cortical labeling protocol. Front. Neurosci. 6, 171, 2012.

34. Rahul S Desikan, Florent Ségonne, Bruce Fischl, Brian T Quinn, Bradford C Dickerson, Deborah Blacker, Randy L Buckner, Anders M Dale, R Paul Maguire, Bradley T Hyman, et al. An automated labeling system for subdividing the human cerebral cortex on MRI scans into gyral based regions of interest. Neuroimage, 31(3):968–980, 2006. doi: 10.1016/j.neuroimage.2006.01.021.

35. Raymond Pomponio, Guray Erus, Mohamad Habes, Jimit Doshi, Dhivya Srinivasan, Eliz-abeth Mamourian, Vishnu Bashyam, Ilya M Nasrallah, Theodore D Satterthwaite, Yong Fan, et al. Harmonization of large MRI datasets for the analysis of brain imaging patterns throughout the lifespan. NeuroImage, 208:116450, 2020. doi: 10.1016/j.neuroimage.2019.116450.

36. Felipe Maia Polo and Renato Vicente. Effective sample size, dimensionality, and generalization in covariate shift adaptation. Neural Computing and Applications, pages 1–13, 2022. doi: 10.1007/s00521-021-06615-1.

37. Ellyn R Butler, Andrew Chen, Rabie Ramadan, Trang T Le, Kosha Ruparel, Tyler M Moore, Theodore D Satterthwaite, Fengqing Zhang, Haochang Shou, Ruben C Gur, et al. Pitfalls in brain age analyses. Technical report, Wiley Online Library, 2021.

38. Ann-Marie G de Lange, Melis Anatürk, Jaroslav Rokicki, Laura KM Han, Katja Franke, Dag Alnæs, Klaus P Ebmeier, Bogdan Draganski, Tobias Kaufmann, Lars T Westlye, et al. Mind the gap: Performance metric evaluation in brain-age prediction. Human Brain Mapping, 2022. doi: 10.1002/hbm.25837.

39. Fabian Pedregosa, Gaël Varoquaux, Alexandre Gramfort, Vincent Michel, Bertrand Thirion, Olivier Grisel, Mathieu Blondel, Peter Prettenhofer, Ron Weiss, Vincent Dubourg, et al. Scikit-learn: Machine learning in Python. the Journal of machine Learning research, 12: 2825–2830, 2011.

40. Adam Paszke, Sam Gross, Soumith Chintala, Gregory Chanan, Edward Yang, Zachary DeVito, Zeming Lin, Alban Desmaison, Luca Antiga, and Adam Lerer. Automatic differentiation in pytorch. 2017.

41. Jes Olesen, André Bes, Robert Kunkel, James W Lance, Giuseppe Nappi, Volker Pfaffenrath, Frank Clifford Rose, Bruce S Schoenberg, Dieter Soyka, Peer Tfelt-Hansen, et al. The international classification of headache disorders, (beta version). Cephalalgia, 33(9): 629–808, 2013. doi: 10.1177/0333102413485658.

42. A.S. Zigmond and R.P. Snaith. The Hospital Anxiety and Depression Scale. Acta Psychiatrica Scandinavica, 67(6):361–370, 1983. doi: 10.1111/j.1600-0447.1983.tb09716.x.

43. Scott M Lundberg and Su-In Lee. A unified approach to interpreting model predictions. Advances in neural information processing systems, 30:4765–4774, 2017.

44. Pedro L Ballester, Jee Su Suh, Natalie CW Ho, Liangbing Liang, Stefanie Hassel, Stephen C Strother, Stephen R Arnott, Luciano Minuzzi, Roberto B Sassi, Raymond W Lam, et al. Gray matter volume drives the brain age gap in schizophrenia: a SHAP study. Schizophrenia, 9 (1):3, 2023. doi: 10.1038/s41537-022-00330-z.

45. Barbara G Tabachnick and Linda S Fidell. Using multivariate statistics. volume 6. Pearson Education Boston, MA, 2013.

46. L. T. Westlye, K. B. Walhovd, A. M. Dale, A. Bjørnerud, P. Due-Tønnessen, A. Engvig, et al. Life-span changes of the human brain white matter: diffusion tensor imaging (DTI) and volumetry. Cerebral cortex, 20(9):2055–2068, 2010. doi: 10.1093/cercor/bhp280.

47. Richard AI Bethlehem, Jakob Seidlitz, Simon R White, Jacob W Vogel, Kevin M Anderson, Chris Adamson, Sophie Adler, George S Alexopoulos, Evdokia Anagnostou, Ariosky Areces-Gonzalez, et al. Brain charts for the human lifespan. Nature, 604(7906):525–533, 2022. doi: 10.1038/s41586-022-04554-y.

48. M Ethan MacDonald and G Bruce Pike. MRI of healthy brain aging: A review. NMR in Biomedicine, 34(9):e4564, 2021. doi: 10.1002/nbm.4564.

49. K. Franke and C. Gaser. Longitudinal changes in individual BrainAGE in healthy aging, mild cognitive impairment, and Alzheimer’s disease. GeroPsych, 25(4):235–245, 2012. doi: 10.1024/1662-9647/a000074.

50. Katja Franke, Christian Gaser, Brad Manor, and Vera Novak. Advanced BrainAGE in older adults with type 2 diabetes mellitus. Frontiers in aging neuroscience, 5:90, 2013. doi: 10.3389/fnagi.2013.00090.

51. James H Cole, Joel Raffel, Tim Friede, Arman Eshaghi, Wallace J Brownlee, Declan Chard, Nicola De Stefano, Christian Enzinger, Lukas Pirpamer, Massimo Filippi, et al. Longitudinal assessment of multiple sclerosis with the brain-age paradigm. Annals of neurology, 88(1): 93–105, 2020. doi: 10.1002/ana.25746.

52. C. Constantinides, L. K. Han, C. Alloza, L. A. Antonucci, C. Arango, R. Ayesa-Arriola, et al. Brain ageing in schizophrenia: evidence from 26 international cohorts via the ENIGMA Schizophrenia consortium. Molecular Psychiatry, 28(3):1201–1209, 2023. doi: 10.1038/s41380-022-01897-w.

53. Didac Vidal-Pineiro, Yunpeng Wang, Stine K Krogsrud, Inge K Amlien, William FC Baaré, David Bartres-Faz, Lars Bertram, Andreas M Brandmaier, Christian A Drevon, Sandra Düzel, et al. Individual variations in ‘brain age’relate to early-life factors more than to longi-tudinal brain change. Elife, 10:e69995, 2021. doi: 10.7554/eLife.69995.

54. J. H. Cole, R. P. Poudel, D. Tsagkrasoulis, M. W. Caan, C. Steves, T. D. Spector, and G. Montana. Predicting brain age with deep learning from raw imaging data results in a reliable and heritable biomarker. NeuroImage, 163:115–124, 2017. doi: 10.1016/j.neuroimage.2017.07.059.

55. Nidhi Gupta, Pushpraj Bhatele, and Pritee Khanna. Glioma detection on brain MRIs using texture and morphological features with ensemble learning. Biomedical Signal Processing and Control, 47:115–125, 2019. doi: 10.1016/j.bspc.2018.06.003.

56. Cherry R Gumiran, Arnel C Fajardo, Ruji P Medina, Minh S Dao, and Betchie E Aguinaldo. Aedes Aegypti Egg Morphological Property and Attribute Determination Based on Computer Vision. In 2022 7th International Conference on Signal and Image Processing (ICSIP), pages 581–585. IEEE, 2022. doi: 10.1109/ICSIP55141.2022.9887255.

57. Peng-Fei Yan, Ling Yan, Ting-Ting Hu, Dong-Dong Xiao, Zhen Zhang, Hong-Yang Zhao, and Jun Feng. The potential value of preoperative MRI texture and shape analysis in grading meningiomas: a preliminary investigation. Translational oncology, 10(4):570–577, 2017. doi: 10.1016/j.tranon.2017.04.006.

58. Saima Rathore, Tamim Niazi, Muhammad Aksam Iftikhar, and Ahmad Chaddad. Glioma grading via analysis of digital pathology images using machine learning. Cancers, 12(3): 578, 2020. doi: 10.3390/cancers12030578.

59. David H Salat, Stephanie Y Lee, AJ Van der Kouwe, Douglas N Greve, Bruce Fischl, and H Diana Rosas. Age-associated alterations in cortical gray and white matter signal intensity and gray to white matter contrast. Neuroimage, 48(1):21–28, 2009. doi: 10.1016/j.neuroimage.2009.06.074.

60. Alvaro Planchuelo-Gomez, David Garcia-Azorin, Angel L Guerrero, Santiago Aja-Fernandez, Margarita Rodriguez, and Rodrigo de Luis-Garcia. Structural connectivity alterations in chronic and episodic migraine: A diffusion magnetic resonance imaging connectomics study. Cephalalgia, 40(4):367–383, 2020. doi: 10.1177/0333102419885392.

61. Álvaro Planchuelo-Gómez, David García-Azorín, Angel L Guerrero, Santiago Aja-Fernández, Margarita Rodríguez, and Rodrigo de Luis-García. Multimodal fusion analysis of structural connectivity and gray matter morphology in migraine. Human Brain Mapping, 42(4):908–921, 2021. doi: 10.1002/hbm.25267.

62. Zhihua Jia and Shengyuan Yu. Grey matter alterations in migraine: a systematic review and meta-analysis. NeuroImage: Clinical, 14:130–140, 2017. doi: 10.1016/j.nicl.2017.01.019.

63. Foucaud du Boisgueheneuc, Richard Levy, Emmanuelle Volle, Magali Seassau, Hughes Duffau, Serge Kinkingnehun, Yves Samson, Sandy Zhang, and Bruno Dubois. Functions of the left superior frontal gyrus in humans: a lesion study. Brain, 129(12):3315–3328, 2006. doi: 10.1093/brain/awl244.

64. Ramon Nogueira, Juan M Abolafia, Jan Drugowitsch, Emili Balaguer-Ballester, Maria V Sanchez-Vives, and Rubén Moreno-Bote. Lateral orbitofrontal cortex anticipates choices and integrates prior with current information. Nature Communications, 8(1):14823, 2017. doi: 10.1038/ncomms14823.

65. Nina Latysheva, Elena Filatova, Diana Osipova, and Alexey B Danilov. Cognitive impairment in chronic migraine: a cross-sectional study in a clinic-based sample. Arquivos de Neuro-Psiquiatria, 78:133–138, 2020. doi: 10.1590/0004-282X20190159.

66. Kerstin Luedtke, Wiebke Starke, and Arne May. Musculoskeletal dysfunction in migraine patients. Cephalalgia, 38(5):865–875, 2018. doi: 10.1177/0333102417716934.

67. James H Cole. Multimodality neuroimaging brain-age in UK biobank: relationship to biomedical, lifestyle, and cognitive factors. Neurobiology of aging, 92:34–42, 2020. doi: https://doi.org/10.1016/j.neurobiolaging.2020.03.014.

68. Mark C Kruit, Mark A van Buchem, Lenore J Launer, Gisela M Terwindt, and Michel D Ferrari. Migraine is associated with an increased risk of deep white matter lesions, subclinical posterior circulation infarcts and brain iron accumulation: the population-based MRI CAM-ERA study. Cephalalgia, 30(2):129–136, 2010. doi: https://doi.org/10.1111/j.1468-2982.2009.01904.x.

